# Estimation of the COVID-19 Average Incubation Time: Systematic Review, Meta-analysis and Sensitivity Analyses

**DOI:** 10.1101/2022.01.17.22269421

**Authors:** Yijia Weng, Grace Y.Yi

## Abstract

**Objectives:** We aim to provide sensible estimates of the average incubation time of COVID-19 by capitalizing available estimates reported in the literature and explore different ways to accommodate heterogeneity involved with the reported studies.

**Methods:** We search through online databases to collect the studies about estimates of the average incubation time and conduct meta-analyses to accommodate heterogeneity of the studies and the publication bias. Cochran’s heterogeneity statistic *Q* and Higgin’s & Thompson’s *I*^2^ statistic are employed. Subgroup analyses are conducted using mixed effects models and publication bias is assessed using the funnel plot and Egger’s test.

**Results:** Using all those reported mean incubation estimates, the average incubation time is estimated to be 6.43 days with a 95% confidence interval (CI) (5.90, 6.96), and using all those reported mean incubation estimates together with those transformed median incubation estimates, the estimated average incubation time is 6.07 days with a 95% CI (5.70,6.45).

**Conclusions:** Providing sensible estimates of the average incubation time for COVID-19 is important yet complex, and the available results vary considerably due to many factors including heterogeneity and publication bias. We take different angles to estimate the mean incubation time, and our analyses provide estimates to range from 5.68 days to 8.30 days.

## 1 Introduction

The coronavirus disease 2019 (COVID-19) has presented tremendous impact on public health as well as economy. Much research has been conducted to understand various clinical characteristics of COVID-19. One interesting question concerns the COVID-19 incubation time which is defined as the time from infection of SARS-CoV-2 to the onset of clinical symptoms (Giesecke, 2017). As the incubation time varies from patient to patient, it is useful to estimate the average incubation time of the population.

Understanding the average incubation time is of great significance for a multitude of reasons. Most obviously, knowing the average incubation time gives us a critical metric in developing strategies for isolation or quarantine. Setting a reasonable quarantine time is directly based on the distribution of incubation times and having a sensible estimate of the average incubation time helps us come up with effective intervention steps. Moreover, in developing epidemic models such as SEIR, the average incubation time is taken as an important parameter to model transmission features of SARS-CoV-2, and different estimates of this parameter may greatly affect the outcomes (Brookmeyer, 2014).

Due to its importance, many studies have been carried out to estimate the average incubation time for COVID-19. However, available studies do not reveal comparable estimates of the mean incubation time, and they vary considerably from 1.8 days in China (Leung, 2020) to 14 days in India (Gupta et al., 2020). Additionally, it is difficult to assess which estimate more reasonably reflects the average incubation time of the population because different studies are carried out for different subjects under different conditions. In this article, we aim to provide synthetic estimates of the average incubation time of COVID-19 by capitalizing on the report estimates in the literature and explore different ways to accommodate heterogeneity involved with the reported studies on COVID-19.

While a number of meta-analyses have offered synthetic estimates, those studies concentrated on early reports which are prior to June 2020, and some of them included a small number of studies. To overcome those limitations, in our analyses here, we conduct a thorough search covering a long study period from January 1, 2020 to May 20, 2021, which allows our study to include more diverse information on estimates of the average incubation estimate. We carry out meta-analyses for those reported mean estimates reported as well as those transformed from estimates of the median incubation time. Subgroup analyses and sensitivity analyses are conducted to investigate heterogeneity among the reported studies and the stability of the produced synthetic estimates of the mean incubation time of COVID-19.

The rest of the manuscript is organized as follows. In Section 2 we present the data collection and extraction methods and the basic characteristics of the data. In Section 3 we describe the general procedures for meta-analyses. In Section 4 we analyze the data and report the results. We conclude the article with discussions presented in the last section.

## 2 Data Collection

### 2.1 Search Strategy and Selection Criteria

We search the articles published between January 1, 2020, and May 20, 2021, through four online databases: *Google Scholar, Web of Science, Scopus*, and *Collabovid*, as well as official websites of journals including *Lancet* and *Journal of American Medical Association*, where *Collabovid* comprises publications from *Elsevier, PubMed, medRxiv, bioRxiv*, and *arXiv*.

We start with an automatic search process using the pairwise combinations of phrases each from one of the following categories: (1). ‘*incubation*’, ‘*incubation period* ‘, and ‘*incubation time*’; (2). ‘*COVID-19* ‘, ‘*SARS-CoV-2* ‘, ‘*2019-nCoV* ‘, ‘*2019nCoV* ‘, and ‘*Novel Coronavirus*’. This process identifies 611 articles. Next, using those keywords, we manually check if the references of the resulting articles should be included in our collection of articles, yielding 17 additional articles. Then we remove 93 duplicated articles from this collection of 628 COVID-19 studies. Further, we examine each study manually by checking first the abstract and then the full text to see whether the study is about the COVID-19 incubation time. The abstract checking process excludes 375 articles. We now further examine the full text for the remaining 160 studies and retain only those studies with the information of the sample size as well as the information on one of the following categories:

a. having an estimate of the *mean* incubation time together with its standard error (SE) or a 95% confidence interval (CI);
b. having an estimate of the *median* incubation time together with a 95% CI, an interquartile range (IQR), or a range.

This step excludes 51 studies for not reporting an estimate of the mean or median incubation time, 2 studies for not reporting dispersion estimates associated with mean or median estimates, and 3 studies for not reporting the sample size. All these procedures finally lead to 104 papers which discuss estimates of the mean or median incubation time for COVID-19.

We summarize this process of gathering the COVID-19 studies about incubation information in Figure 1, which is prepared using the flow chart template developed for systematic review and meta-analysis, available at the website www.prisma-statement.org.

**Figure 1:**
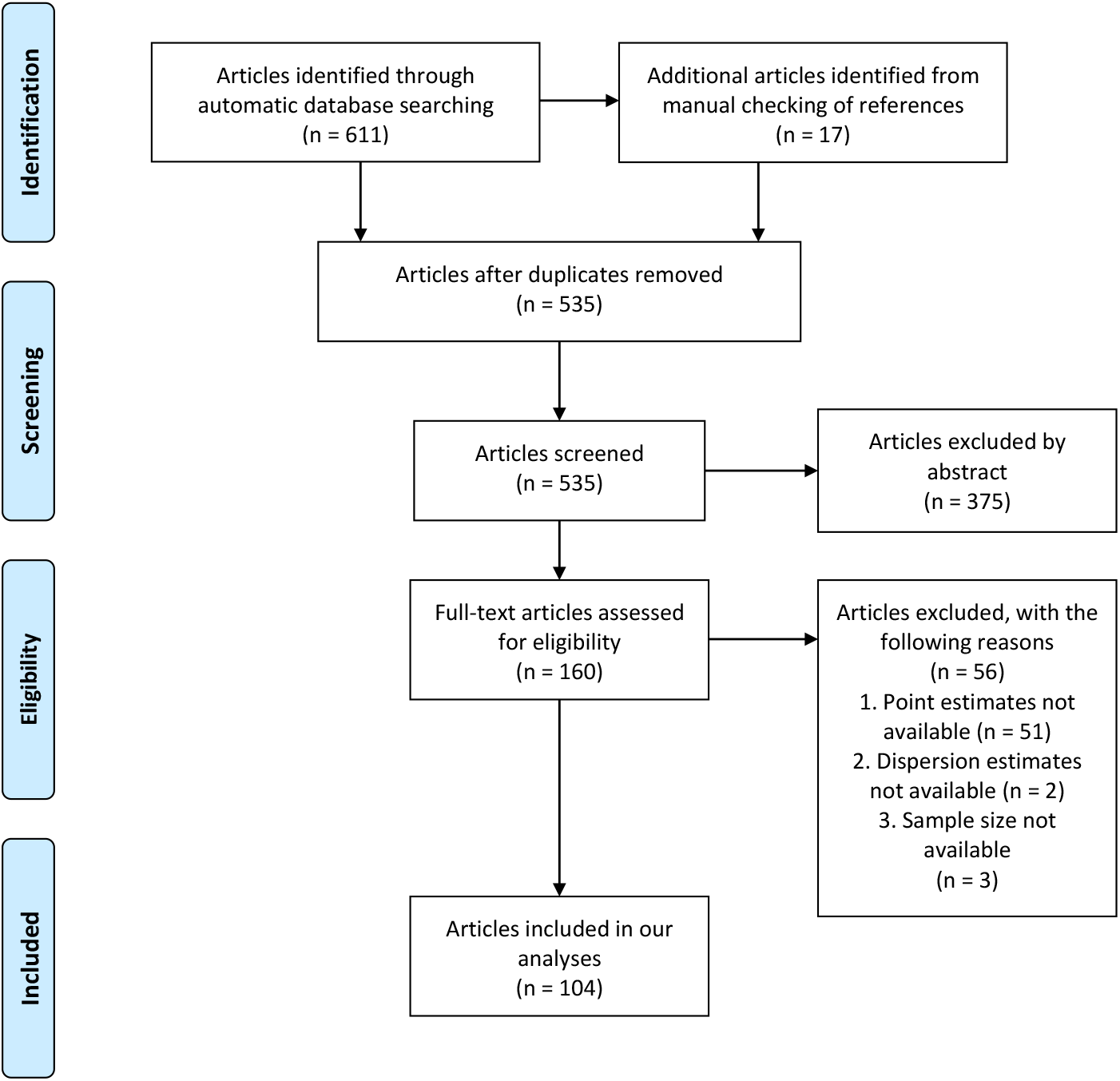
Flow diagram for gathering studies about estimation of the mean or median COVID-19 incubation time

### 2.2 Data Extraction

We now report those 104 papers searched in Section 2.1 by displaying the information about the last name of the first author, the study period, the region of study subjects, and the methodology, together with the sample size, the estimate of the mean or median COVID-19 incubation time, and the associated standard error reported in the article or converted by us from using the reported 95% CI.

In Figure 2 we further display the information of those selected 104 papers, in which 69 (*N*_1_) studies merely report the information about estimates of the mean incubation time and 35 (*N*_2_) articles report only the information about estimates of the median incubation time. Those 69 (*N*_1_) papers can be further grouped as 16 (*N*_11_) papers containing meta-analysis results each derived from multiple studies and 53 (*N*_12_) papers each reporting results obtained from a single study, where in those 16 (*N*_11_) papers, 1 (*N*_111_) paper reports two estimates with one synthetic estimate derived from multiple studies using the meta-analysis method and the other estimate obtained from a single new study, and 15 (*N*_112_) papers each reports a single estimate obtained from meta-analysis. In those 53 (*N*_12_) papers, 1 (*N*_121_) paper reports three mean estimates, 3 (*N*_122_) papers each reports two mean estimates, and 49 (*N*_123_) papers each reports a single estimate. Those 35 (*N*_2_) papers consist of 1 (*N*_21_) paper reporting two median estimates and 34 (*N*_22_) papers each reporting a single median estimate.

**Figure 2:**
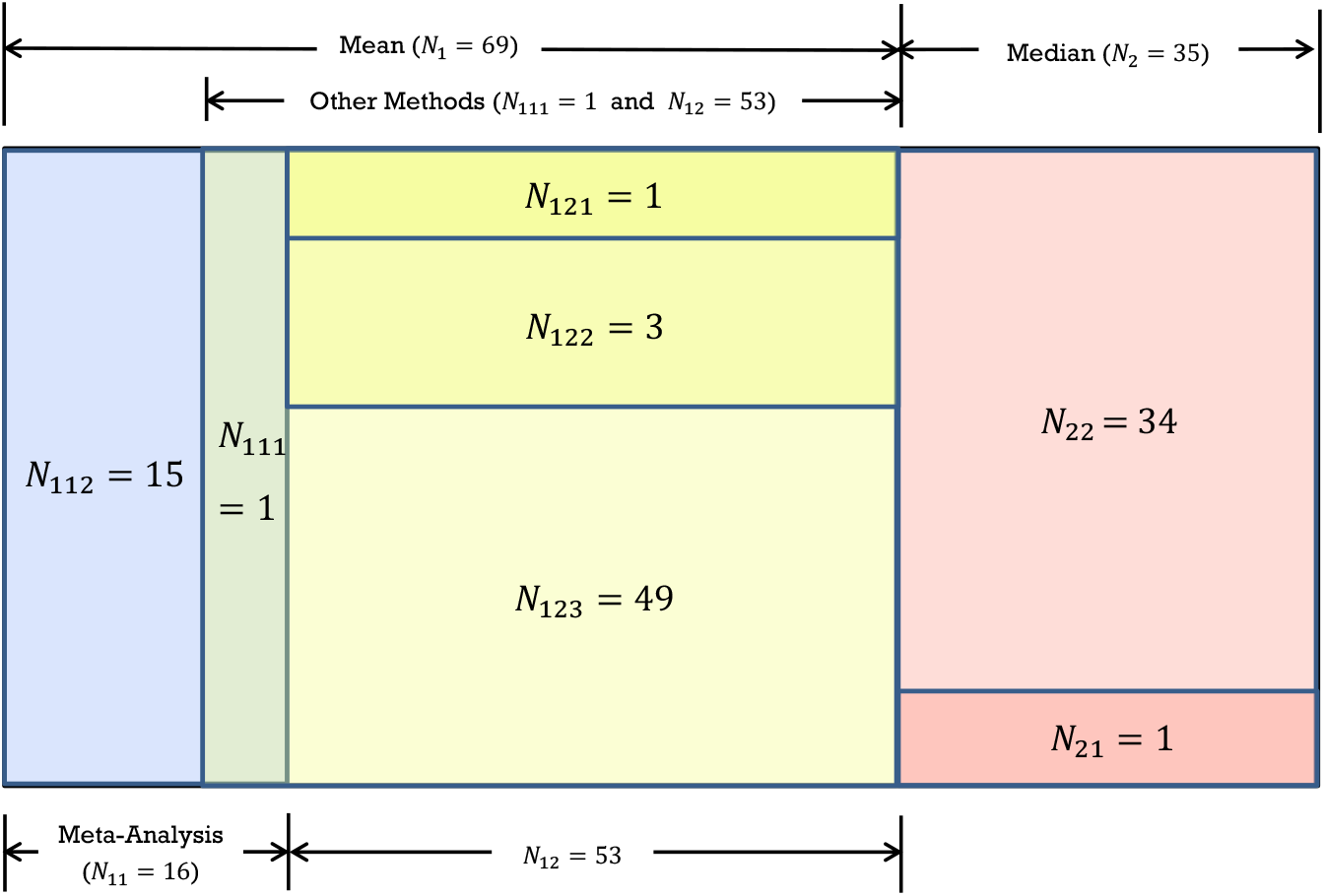
Nested Structures in the collected papers

With the summary of the number of papers and of estimates shown in Table 1, we now provide more detailed information for the 104 papers in Tables 2-4. To be specific, 16 (*N*_11_) papers with 16 (*N*_112_ · 1 + *N*_111_ · 1) mean incubation estimates using meta-analysis methods are displayed in Table 2, 54 (*N*_111_ + *N*_12_) papers with 59 (*N*_111_ · 1 + *N*_121_ · 3 + *N*_122_ · 2 + *N*_123_ · 1) mean incubation estimates with methods other than meta-analysis are displayed in Table 3, and 35 (*N*_2_) papers with 36 (*N*_21_ · 2 + *N*_22_ · 1) median incubation estimates are displayed in Table 4.

**Table 1:**
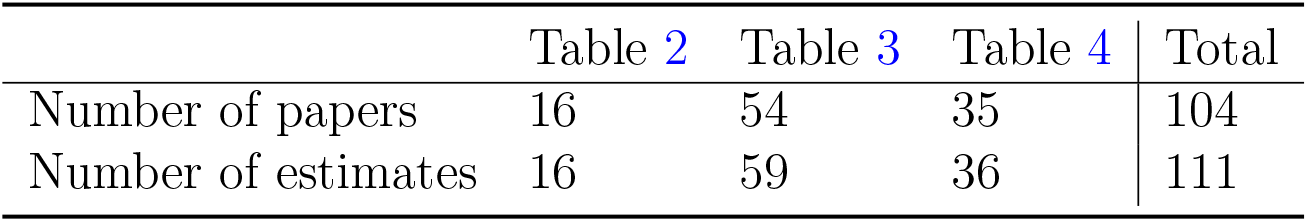
The number of papers and estimates reported in Tables 2-4

**Table 2:** A summary of 16 papers reporting meta-analysis results about estimation of the mean incubation time

| Author | Period | Region | Methodology | Sample Size | Mean | SD |
| --- | --- | --- | --- | --- | --- | --- |
| He et al. | Up to 24 Feb 2020 | Worldwide | Meta-analysis | 5 studies | 5.08 | 0.16 |
| Li et al. | 1 Jan-6 Apr 2020 | Worldwide | Meta-analysis | 7 studies (746) | 5.30 | 0.38 |
| Quesada et al. | 1 Jan-21 March | Worldwide | Meta-analysis | 7 studies (792) | 5.60 | 0.26 |
| Zhang et al. | 1 Jan-24 Feb 2020 | Worldwide | Meta-analysis | 11 studies (3607) | 5.34 | 0.54 |
| Alene et al. | Up to 31 Mar 2020 | Worldwide | Meta-analysis | 14 studies (1458) | 6.50 | 0.31 |
| Rai et al. | Up tp 31 Mar 2020 | Worldwide | Meta-analysis | 15 studies | 5.74 | 0.27 |
| Wassie et al. | Up to 2 May 2020 | Worldwide | Meta-analysis | 18 studies (22595) | 5.70 | 0.33 |
| McAloon et al. | Up to 27 Feb 2020 | Worldwide | Meta-analysis(only log normal) | 24 studies (1357) | 5.80 | 0.43 |
| Dhouib et al. | Dec 2019-Mar 2020 | China | Meta-analysis | 42 studies | 6.20 | 0.41 |
| Zhang et al.<sup>†</sup> | Up to 8 May 2020 | Worldwide | Meta-analysis | 42 studies (13272) | 6.25 | 0.26 |
| Khalili et al. | 1 Dec 2019-11 Mar 2020 | Worldwide | Meta-analysis | 43 studies | 5.68 | 0.46 |
| Wang et al. | 23 Jan-20 Mar 2020 | Worldwide | Meta-analysis | 47 studies | 5.44 | 0.26 |
| Pormohammad et al. | Up tp0 26 Apr 2020 | Worldwide | Meta-analysis | 53 studies (12609) | 6.40 | 0.31 |
| Wei et al. | 1 Dec 2019-24 Apr 2020 | Worldwide | Meta-analysis(only log normal) | 56 studies (4095) | 5.80 | 0.23 |
| Banka et al. | 1 Jan-27 Jul 2020 | Worldwide | Meta-analysis(Gamma) | 64 studies (45151) | 6.71 | 2.91 |
| Elias et al. | 1 Jan 2020-10 Jan 2021 | Mainly in Asia | Meta-analysis | 99 studies | 6.38 | 0.26 |

<sup>†</sup> This paper ( $N_{111}$ ) reports one synthetic mean incubation estimate derived from multiple studies using meta-analysis and one mean incubation estimate obtained from a single sample.
The number in brackets under the heading “Sample Size” represent number of total sample size within all meta-analyses. (if the author provided.)

**Table 3:**
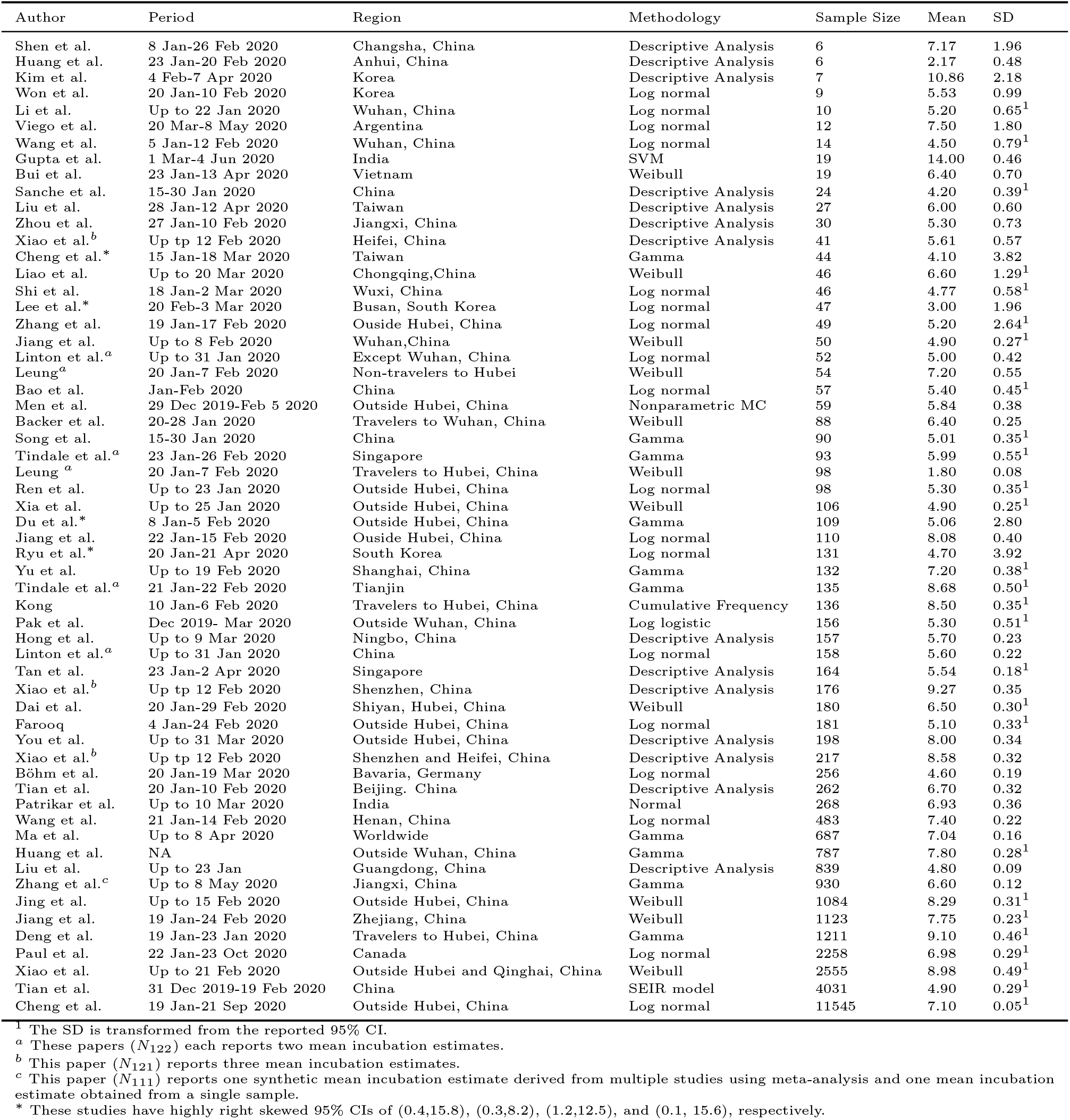
A summary of 59 estimates of the mean incubation time from 54 papers

| Author | Period | Region | Methodology | Sample Size | Mean | SD |
| --- | --- | --- | --- | --- | --- | --- |
| Shen et al. | 8 Jan-26 Feb 2020 | Changsha, China | Descriptive Analysis | 6 | 7.17 | 1.96 |
| Huang et al. | 23 Jan-20 Feb 2020 | Anhui, China | Descriptive Analysis | 6 | 2.17 | 0.48 |
| Kim et al. | 4 Feb-7 Apr 2020 | Korea | Descriptive Analysis | 7 | 10.86 | 2.18 |
| Won et al. | 20 Jan-10 Feb 2020 | Korea | Log normal | 9 | 5.53 | 0.99 |
| Li et al. | Up to 22 Jan 2020 | Wuhan, China | Log normal | 10 | 5.20 | 0.65<sup>1</sup> |
| Viego et al. | 20 Mar-8 May 2020 | Argentina | Log normal | 12 | 7.50 | 1.80 |
| Wang et al. | 5 Jan-12 Feb 2020 | Wuhan, China | Log normal | 14 | 4.50 | 0.79<sup>1</sup> |
| Gupta et al. | 1 Mar-4 Jun 2020 | India | SVM | 19 | 14.00 | 0.46 |
| Bui et al. | 23 Jan-13 Apr 2020 | Vietnam | Weibull | 19 | 6.40 | 0.70 |
| Sanche et al. | 15-30 Jan 2020 | China | Descriptive Analysis | 24 | 4.20 | 0.39<sup>1</sup> |
| Liu et al. | 28 Jan-12 Apr 2020 | Taiwan | Descriptive Analysis | 27 | 6.00 | 0.60 |
| Zhou et al. | 27 Jan-10 Feb 2020 | Jiangxi, China | Descriptive Analysis | 30 | 5.30 | 0.73 |
| Xiao et al.<sup>b</sup> | Up tp 12 Feb 2020 | Heifei, China | Descriptive Analysis | 41 | 5.61 | 0.57 |
| Cheng et al.* | 15 Jan-18 Mar 2020 | Taiwan | Gamma | 44 | 4.10 | 3.82 |
| Liao et al. | Up to 20 Mar 2020 | Chongqing, China | Weibull | 46 | 6.60 | 1.29<sup>1</sup> |
| Shi et al. | 18 Jan-2 Mar 2020 | Wuxi, China | Log normal | 46 | 4.77 | 0.58<sup>1</sup> |
| Lee et al.* | 20 Feb-3 Mar 2020 | Busan, South Korea | Log normal | 47 | 3.00 | 1.96 |
| Zhang et al. | 19 Jan-17 Feb 2020 | Ouside Hubei, China | Log normal | 49 | 5.20 | 2.64<sup>1</sup> |
| Jiang et al. | Up to 8 Feb 2020 | Wuhan, China | Weibull | 50 | 4.90 | 0.27<sup>1</sup> |
| Linton et al.<sup>a</sup> | Up to 31 Jan 2020 | Except Wuhan, China | Log normal | 52 | 5.00 | 0.42 |
| Leung<sup>a</sup> | 20 Jan-7 Feb 2020 | Non-travelers to Hubei | Weibull | 54 | 7.20 | 0.55 |
| Bao et al. | Jan-Feb 2020 | China | Log normal | 57 | 5.40 | 0.45<sup>1</sup> |
| Men et al. | 29 Dec 2019-Feb 5 2020 | Outside Hubei, China | Nonparametric MC | 59 | 5.84 | 0.38 |
| Backer et al. | 20-28 Jan 2020 | Travelers to Wuhan, China | Weibull | 88 | 6.40 | 0.25 |
| Song et al. | 15-30 Jan 2020 | China | Gamma | 90 | 5.01 | 0.35<sup>1</sup> |
| Tindale et al.<sup>a</sup> | 23 Jan-26 Feb 2020 | Singapore | Gamma | 93 | 5.99 | 0.55<sup>1</sup> |
| Leung<sup>a</sup> | 20 Jan-7 Feb 2020 | Travelers to Hubei, China | Weibull | 98 | 1.80 | 0.08 |
| Ren et al. | Up to 23 Jan 2020 | Outside Hubei, China | Log normal | 98 | 5.30 | 0.35<sup>1</sup> |
| Xia et al. | Up to 25 Jan 2020 | Outside Hubei, China | Weibull | 106 | 4.90 | 0.25<sup>1</sup> |
| Du et al.* | 8 Jan-5 Feb 2020 | Outside Hubei, China | Gamma | 109 | 5.06 | 2.80 |
| Jiang et al. | 22 Jan-15 Feb 2020 | Ouside Hubei, China | Log normal | 110 | 8.08 | 0.40 |
| Ryu et al.* | 20 Jan-21 Apr 2020 | South Korea | Log normal | 131 | 4.70 | 3.92 |
| Yu et al. | Up to 19 Feb 2020 | Shanghai, China | Gamma | 132 | 7.20 | 0.38<sup>1</sup> |
| Tindale et al.<sup>a</sup> | 21 Jan-22 Feb 2020 | Tianjin | Gamma | 135 | 8.68 | 0.50<sup>1</sup> |
| Kong | 10 Jan-6 Feb 2020 | Travelers to Hubei, China | Cumulative Frequency | 136 | 8.50 | 0.35<sup>1</sup> |
| Pak et al. | Dec 2019- Mar 2020 | Outside Wuhan, China | Log logistic | 156 | 5.30 | 0.51<sup>1</sup> |
| Hong et al. | Up to 9 Mar 2020 | Ningbo, China | Descriptive Analysis | 157 | 5.70 | 0.23 |
| Linton et al.<sup>a</sup> | Up to 31 Jan 2020 | China | Log normal | 158 | 5.60 | 0.22 |
| Tan et al. | 23 Jan-2 Apr 2020 | Singapore | Descriptive Analysis | 164 | 5.54 | 0.18<sup>1</sup> |
| Xiao et al.<sup>b</sup> | Up tp 12 Feb 2020 | Shenzhen, China | Descriptive Analysis | 176 | 9.27 | 0.35 |
| Dai et al. | 20 Jan-29 Feb 2020 | Shiyan, Hubei, China | Weibull | 180 | 6.50 | 0.30<sup>1</sup> |
| Farooq | 4 Jan-24 Feb 2020 | Outside Hubei, China | Log normal | 181 | 5.10 | 0.33<sup>1</sup> |
| You et al. | Up to 31 Mar 2020 | Outside Hubei, China | Descriptive Analysis | 198 | 8.00 | 0.34 |
| Xiao et al.<sup>b</sup> | Up tp 12 Feb 2020 | Shenzhen and Heifei, China | Descriptive Analysis | 217 | 8.58 | 0.32 |
| Böhm et al. | 20 Jan-19 Mar 2020 | Bavaria, Germany | Log normal | 256 | 4.60 | 0.19 |
| Tian et al. | 20 Jan-10 Feb 2020 | Beijing, China | Descriptive Analysis | 262 | 6.70 | 0.32 |
| Patrikar et al. | Up to 10 Mar 2020 | India | Normal | 268 | 6.93 | 0.36 |
| Wang et al. | 21 Jan-14 Feb 2020 | Henan, China | Log normal | 483 | 7.40 | 0.22 |
| Ma et al. | Up to 8 Apr 2020 | Worldwide | Gamma | 687 | 7.04 | 0.16 |
| Huang et al. | NA | Outside Wuhan, China | Gamma | 787 | 7.80 | 0.28<sup>1</sup> |
| Liu et al. | Up to 23 Jan | Guangdong, China | Descriptive Analysis | 839 | 4.80 | 0.09 |
| Zhang et al.<sup>c</sup> | Up to 8 May 2020 | Jiangxi, China | Gamma | 930 | 6.60 | 0.12 |
| Jing et al. | Up to 15 Feb 2020 | Outside Hubei, China | Weibull | 1084 | 8.29 | 0.31<sup>1</sup> |
| Jiang et al. | 19 Jan-24 Feb 2020 | Zhejiang, China | Weibull | 1123 | 7.75 | 0.23<sup>1</sup> |
| Deng et al. | 19 Jan-23 Jan 2020 | Travelers to Hubei, China | Gamma | 1211 | 9.10 | 0.46<sup>1</sup> |
| Paul et al. | 22 Jan-23 Oct 2020 | Canada | Log normal | 2258 | 6.98 | 0.29<sup>1</sup> |
| Xiao et al. | Up to 21 Feb 2020 | Outside Hubei and Qinghai, China | Weibull | 2555 | 8.98 | 0.49<sup>1</sup> |
| Tian et al. | 31 Dec 2019-19 Feb 2020 | China | SEIR model | 4031 | 4.90 | 0.29<sup>1</sup> |
| Cheng et al. | 19 Jan-21 Sep 2020 | Outside Hubei, China | Log normal | 11545 | 7.10 | 0.05<sup>1</sup> |

<sup>1</sup> The SD is transformed from the reported 95% CI.
<sup>a</sup> These papers ( $N_{122}$ ) each reports two mean incubation estimates.
<sup>b</sup> This paper ( $N_{121}$ ) reports three mean incubation estimates.
<sup>c</sup> This paper ( $N_{111}$ ) reports one synthetic mean incubation estimate derived from multiple studies using meta-analysis and one mean incubation estimate obtained from a single sample.
\* These studies have highly right skewed 95% CIs of (0.4,15.8), (0.3,8.2), (1.2,12.5), and (0.1, 15.6), respectively.

**Table 4:**
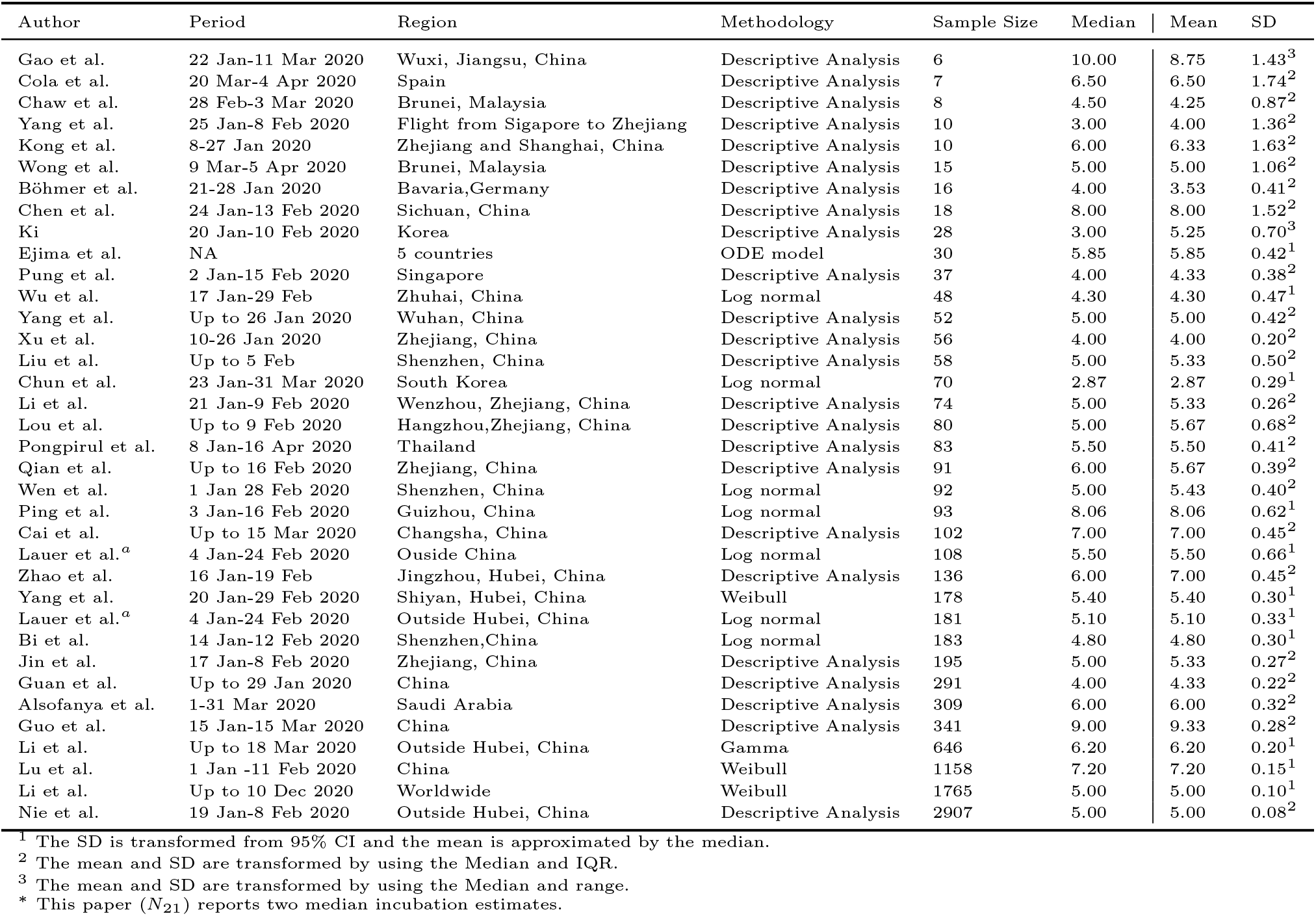
A summary of 36 estimates about the median incubation time from 35 papers, together with the derived entries reported in the last two columns

In Tables 2 and 3, only information on estimates of the mean incubation time is included; and in Table 4, we show 35 studies with 36 reported estimates of median incubation time, together with the computed estimates of the mean and standard deviation (SD) using the methods described in Section 4.3 and Section S2 of the Supplementary Material.

Among the papers on meta-analysis reported in Table 2, the size of studies in each paper varies from 5 to 99, and the estimates (in days) of the mean incubation time range from 5.08 (He et al., 2020) to 6.71 (Banka and Comiskey, 2020). Of all those 16 meta-analyses, 14 are conducted for worldwide studies, one is for patients in China, and one is for patients in Asia. In terms of the distributional assumption for the incubation time, 2 papers assume a log-normal distribution, 1 paper assumes a gamma distribution, and 13 papers make other assumptions.

Among the studies reported in Table 3, the sample size varies from 6 to 11,545, and the estimates of the mean incubation time range from 1.8 to 14 days. Forty-one (74.55%) studies are conducted inside China, in which 9 (16.36%) estimates are obtained from study subjects inside Hubei province, China. In terms of the methodology, 14 (25.45%) analyses are descriptive, 37 (67.27%) studies are derived from parametric models, and the rest are from non-parametric models. For those studies not reporting the standard deviation (SD) but reporting a 95% confidence interval (CI) of the mean incubation time, we use the length of the 95% confidence interval *L* to estimate SD:

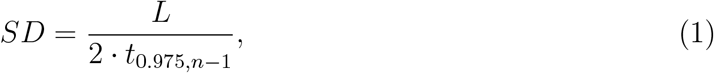

where *t*_0.975,*n*−1_ is the 97.5th percentile of the student’s *t* distribution with (*n* − 1) degrees of freedom, and *n* is the sample size of the study (Higgins et al., 2019). Reported and estimated SDs are shown in the last column of Table 3.

Among the studies reported in Table 4, the reported sample size varies from 6 to 2907, and the estimates of the median incubation time range from 2.87 to 10.00 days. The computed estimates of the mean incubation time vary from 2.87 to 9.33 days. Twenty-three (64.86%) studies are conducted inside China, in which 4 (11.11%) of them are inside Hubei province, China. In terms of the methodology, 24 (66.67%) analyses are descriptive, 11 (30.56%) studies are derived from parametric models, and one study (2.78%) uses non-parametric models. To estimate the SD, we apply (1) to those studies with 95% CI reported. For studies with only interquartile range (IQR) or range, we transform those quantities to obtain estimates of the mean incubation time and SD (Wan et al., 2014) using the formulas displayed in S2 of the Supplementary Materials.

## 3 Meta-Analysis: Models and Procedures

Our objective is to provide sensible estimates of the mean incubation time of COVID-19 by capitalizing on the results reported in the literature with the study heterogeneity taken into account. We particularly examine the studies reported in Table 3 or 4 with the meta-analysis method under random effects or fixed effect models. Before reporting the analysis results, in this section we review the general procedure of meta-analysis.

### 3.1 Random Effects Model and Fixed Effect Model

Let *μ* denote the true mean incubation time for the population, and suppose that *K* independent studies are available to report an estimate of *μ*. For *i* = 1, …, *K*, let *y*_*i*_ denote the estimate of *μ*, and let *σ*_*i*_ denote the associated standard error.

In conducting the meta-analysis under the fixed effect model (Jackson and White, 2018), we use a pooled average of the estimates from the *K* studies to obtain a synthetic estimate of *μ*, given by

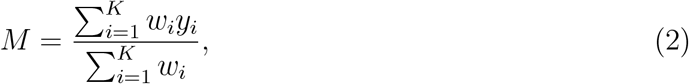

and the associated variance for *M* is calculated as:

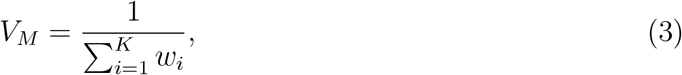

where !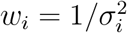 is the weight for study *i*.

Similarly, when conducting meta-analysis under the random effects model, a synthetic estimate of *μ* and its associated variance, denoted *M* ^*\**^ and *V*_*M*_*\**, respectively, are still given by (2) and (3), except that the weight *w*_*i*_ for study *i* is now replaced by 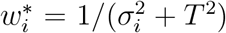 (Borenstein et al., 2009, pp.77-87), where 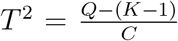, with 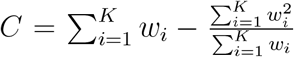 and 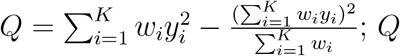; *Q* is also Cochran’s heterogeneity statistic.

### 3.2 Heterogeneity Test

To assess the heterogeneity among different studies, we are interested in testing the null hypothesis:

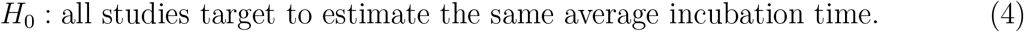

One approach is to use Cochran’s heterogeneity statistic *Q* to calculate the *p*-value, *P* (*χ*^2^(*K*− 1) *> Q*), with *χ*^2^(*K* − 1) representing a random variable following the *χ*^2^ distribution of (*K* − 1) degrees of freedom (Cochran, 1954).

Another useful approach is based on the so-called *I*^2^ statistic (Higgins and Thompson, 2002), defined as

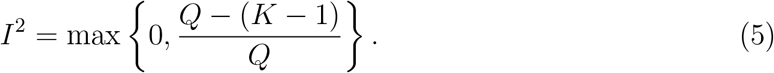

If *I*^2^ *>* 50%, a random effect model is preferred; otherwise, a fixed effect model is suggested. Moreover, substantial heterogeneity is revealed if *I*^2^ *>* 75% (Higgins et al., 2003).

Both *Q* and *I*^2^ statistics do not depend on the scale of measurements, but their performance differently depends on *K*. The *Q* statistic is more sensitive to small values of *K* than the *I*^2^ statistic does. When *K* is smaller than 10, the *Q* statistic may not perform reliably. *I*^2^ explores the between-study variance in a relative scale whereas *Q* statistic explains the variance in the absolute scale.

### 3.3 Forest Plot

To visualize the results from meta-analysis in contrast to the results reported by individual studies, one may employ the forest plot (Lalkhen and McCluskey, 2008). Here we use the package *meta* (Balduzzi et al., 2019) in R version 4.1.0 to construct forest plots, which displays the information for each study, including the last name of the first author and the estimate of the mean incubation time with a 95% CI, together with the pooled average incubation estimate and a 95% CI as well as the *I*^2^ statistics, *Q* statistics, and its *p*-value.

### 3.4 Subgroup Analyses

If the random effects model is suggested by the test in Section 3.2, one may further investigate the source of heterogeneity among the *K* studies by conducting subgroup analyses with different groupings introduced (Borenstein and Higgins, 2013). The mixed effects model is taken to reflect the differences in the mean incubation times between subgroups, where the subgroups are assumed to be fixed and within each subgroup, the true effect size (i.e., the mean incubation time here) varies across different studies. The method of testing for the between-group difference is conducted by first taking the pooled results of random effects model within each subgroup as single estimates and then applying the Cochran’s heterogeneity statistic *Q* described in Section 3.1 (Borenstein and Higgins, 2013).

To be specific, suppose we divide all the studies into *S* subgroups. For *i* = 1, …, *S*, we use the procedures for obtaining 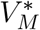 and *M* ^*\**^ under the random effects model described in Section 3.1 to calculate the estimate of the mean incubation time and the associated standard error for subgroup *i*, denoted *s*_*i*_ and *τ*_*i*_, respectively. By replacing *y*_*i*_ with *s*_*i*_ and the *σ*_*i*_ with *τ*_*i*_ in the *Q* statistic in Section 3.1, we obtain the Cochran’s heterogeneity statistic, denoted *Q*^*\**^, for heterogeneity among the *S* subgroups. Then we apply the test procedure described in Section 3.2 to assess if the mean incubation time estimates differ significantly among the subgroups.

### 3.5 Risk of Bias Assessment

To evaluate the quality of the studies, it is important to assess the risk of bias, defined as the systematic error or deviation from the truth (Viswanathan et al., 2012). We adapt the risk of bias tool developed by Hoy et al. (2012) and use a 10-point checklist to assess the risk of bias for each study. In particular, in items 9 and 10 of the checklist of Hoy et al. (2012) which are about disease prevalence, we change the descriptions to reflect the information on the COVID-19 incubation times as suggested by Quesada et al. (2021). The checklist includes both external and internal bias assessments related to the sampling method, data collection, case definition, the validity of methodology, and reporting bias as suggested by Wassie et al. (2020). The answer to each question in the list is ranked as low risk (1 point) or high risk (0 point). A total score over 8 is regarded as an overall indication of low risk of bias, a total score below 5 indicates high risk of bias, and otherwise moderate risk of bias. The details of the checklist are displayed in Section S1 of the Supplementary Material.

To display results of assessing the risk of bias, one may use the function *rob*.*summary()* in package *dmetar* (Harrer et al., 2019) in R version 4.1.0 which outputs two summary tables. One summary table reports the proportion of studies with high/low risk of bias for each checklist question, and the other table, called *RevMan risk of bias table* (Higgins and Thomas, 2011), displays a more detailed view by showing the risk of bias results associated with each study for each question, where the rows correspond to the risk assessment items and the columns refer to the studies; in the display, color red or blue is used to show high and low risk of bias, respectively.

### 3.6 Publication Bias

To understand the results produced by the meta-analysis, we assess potential publication bias incurred in individual studies. To this end, we use the funnel plot and Egger’s test (Egger et al., 1997).

The funnel plot displays the standard error against the effect size for each study. If publication bias is present, the funnel would look asymmetrical. To measure asymmetry of the funnel plot, one may employ Egger’s test which involves a linear regression equation (Egger et al., 1997):

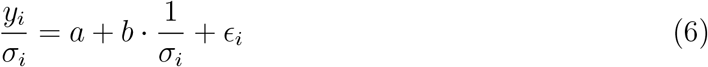

for *i* = 1,.., *K*, where *a* and *b* are the intercept and slope, and *E*_*i*_ is the noise term with mean zero. Then assessing no publication bias is reflected by the null hypothesis:

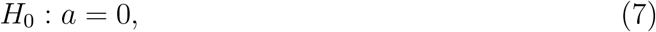

and the test statistic is calculated as:

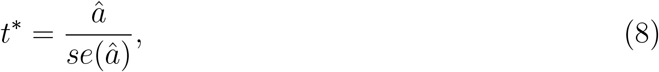

where *â* refers to the estimates of *a* and *se*(*â*) is the associated standard error by applying the least squares method to fit model (6) to the data {(*y*_*i*_,*σ*_*i*_): *i* = 1, …, *K*}. Then the *p*-value of testing (7) is given by 2 *· P* (*t*(*K* − 2) *>* |*t*^*\**^|), where *t*(*K* − 2) represents a random variable having the *t* distribution with (*K* − 2) degrees of freedom. A small *p*-value indicates the presence of the publication bias.

## 4 Data Analysis

Now we apply the procedures described in Section 3 to analyze the data described in Section 2. We carry out four different analyses. *Analysis 1* is conducted to those studies with the information about estimates of the mean incubation time only, whereas *Analysis 2* are based on the studies with the information about estimates of the median incubation time. *Analysis 3* combines the studies in both *Analyses 1* and *2*, where a transformation described in Section S2 of the Supplementary Material is used to convert the estimates of the median incubation time to those of the mean incubation time. *Analysis 4* is conducted to those studies which merely report meta-analysis results.

Further, using the methods in Section 3.4, we perform subgroup analyses for different regions (China and outside China, Hubei and outside Hubei), different methodologies (parametric models, non-parametric models, and descriptive analysis), and different bias levels (low risk, moderate risk, and high risk). These analyses allow us to evaluate estimates with certain heterogeneities controlled.

### 4.1 Assessing Risk of Bias and Publication Bias

Risk of bias assessment and publication bias assessment are conducted using methods described in Sections 3.5 and 3.6, respectively. Figure 3 shows an overall summary for all the 95 studies in Tables 3 and 4 with those 16 studies about meta-analysis in Table 2 excluded, and Figure 4 displays the RevMan risk of bias table, where the risk status (high or low) for 10 questions in the checklist and 95 studies is displayed by rows and columns, respectively. Overall, 5.26% of the studies are of low risk of bias, 43.16% are of moderate risk of bias, and 51.58% are of high risk of bias. There is no evidence of publication bias for *Analyses 1-4* ; details of each test are provided in Sections 4.2-4.5.

**Figure 3:**
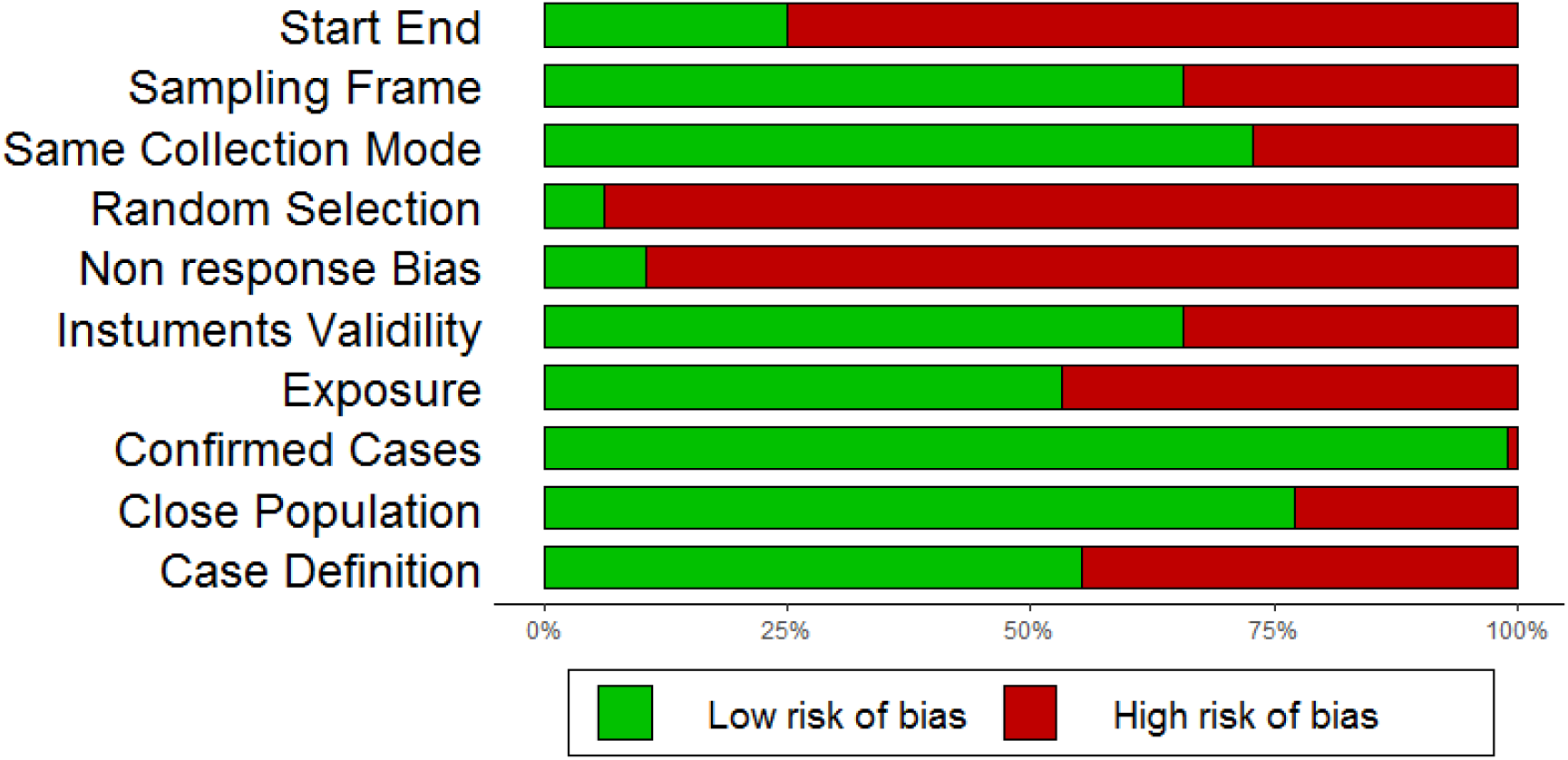
Summary of risk of bias

**Figure 4:**
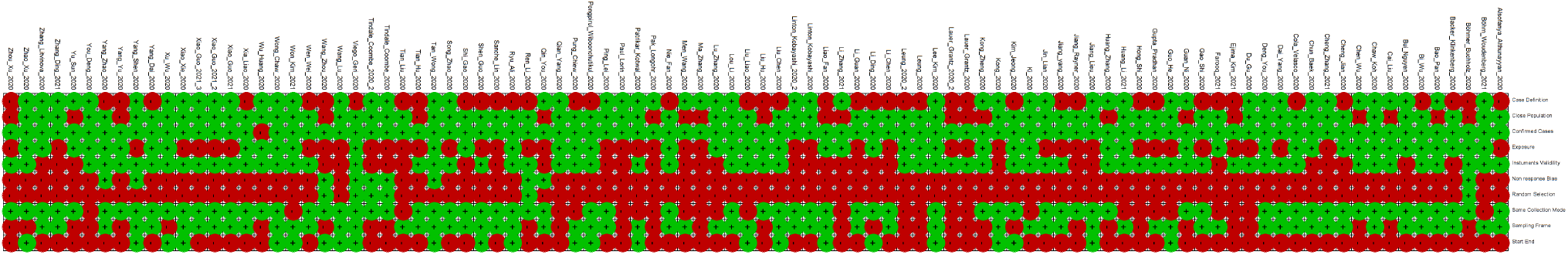
RevMan risk of bias table

### 4.2 Results of Analysis 1

Table 3 contains 4 studies (marked as an asterisk) with highly right screwed 95% CIs, in the sense that the mean is a lot closer to the lower bound than the upper bound, suggesting that the derived standard deviations using (1) are unreliable. Thus, we exclude those studies from the 59 studies summarized in Table 3 and then apply the test procedures described in Section 3.2 to the remaining 55 studies. The *p*-value for Cochran’s test is less than 0.01 and *I*^2^ = 99%, suggesting that the random effects model is preferred when conducting meta-analysis. Figure 5 displays the forest plot of the meta-analysis, showing that the pooled mean incubation estimate for *Analysis 1* is 6.43 days with a 95% CI (5.90, 6.96). By applying the method in Section 3.6, we obtain the *p*-value 0.33 for the Egger’s test, suggesting no evidence for the presence of asymmetry in the funnel plot, displayed in Figure 6.

**Figure 5:**
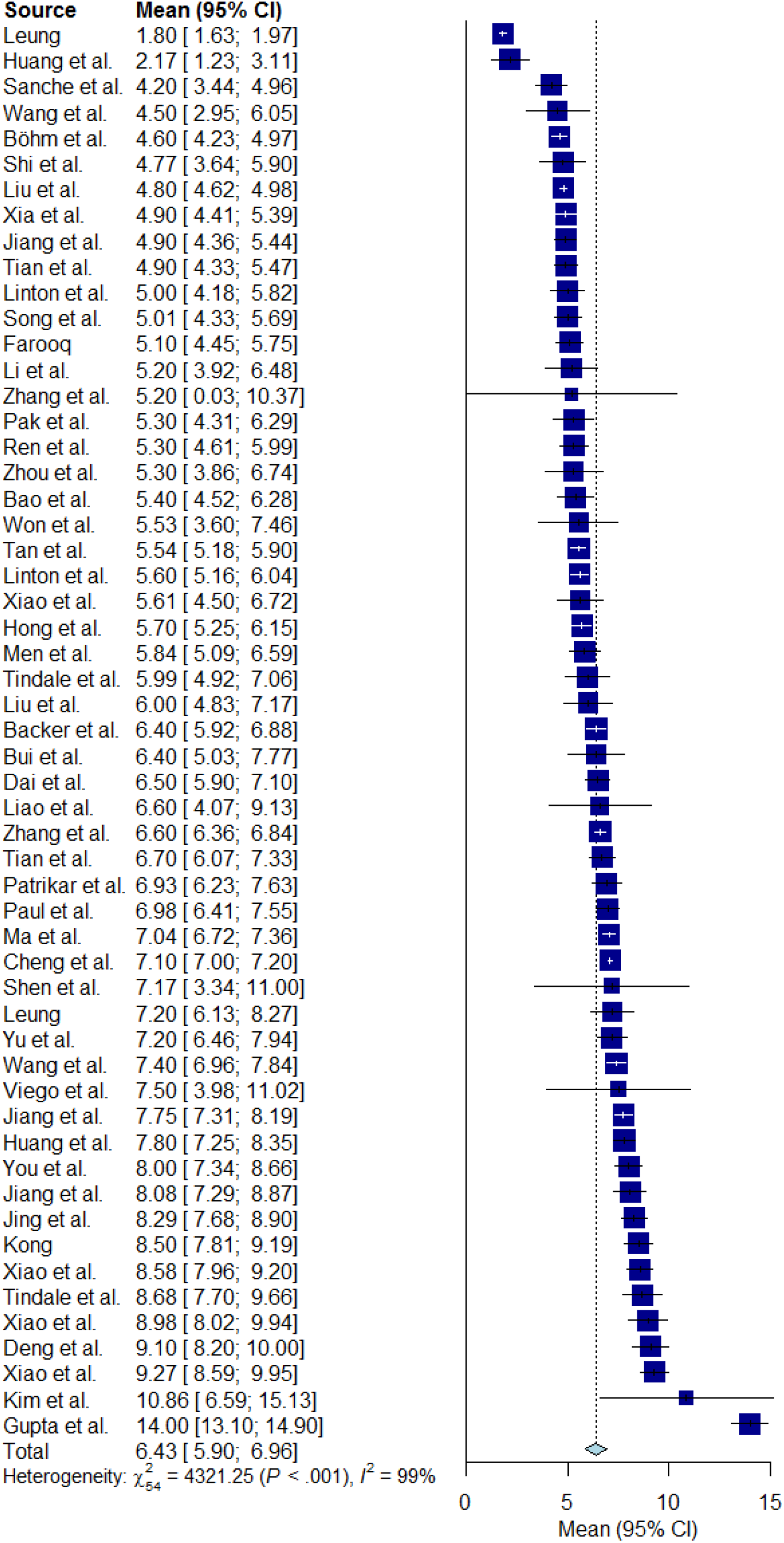
Forest plot for Analysis 1

**Figure 6:**
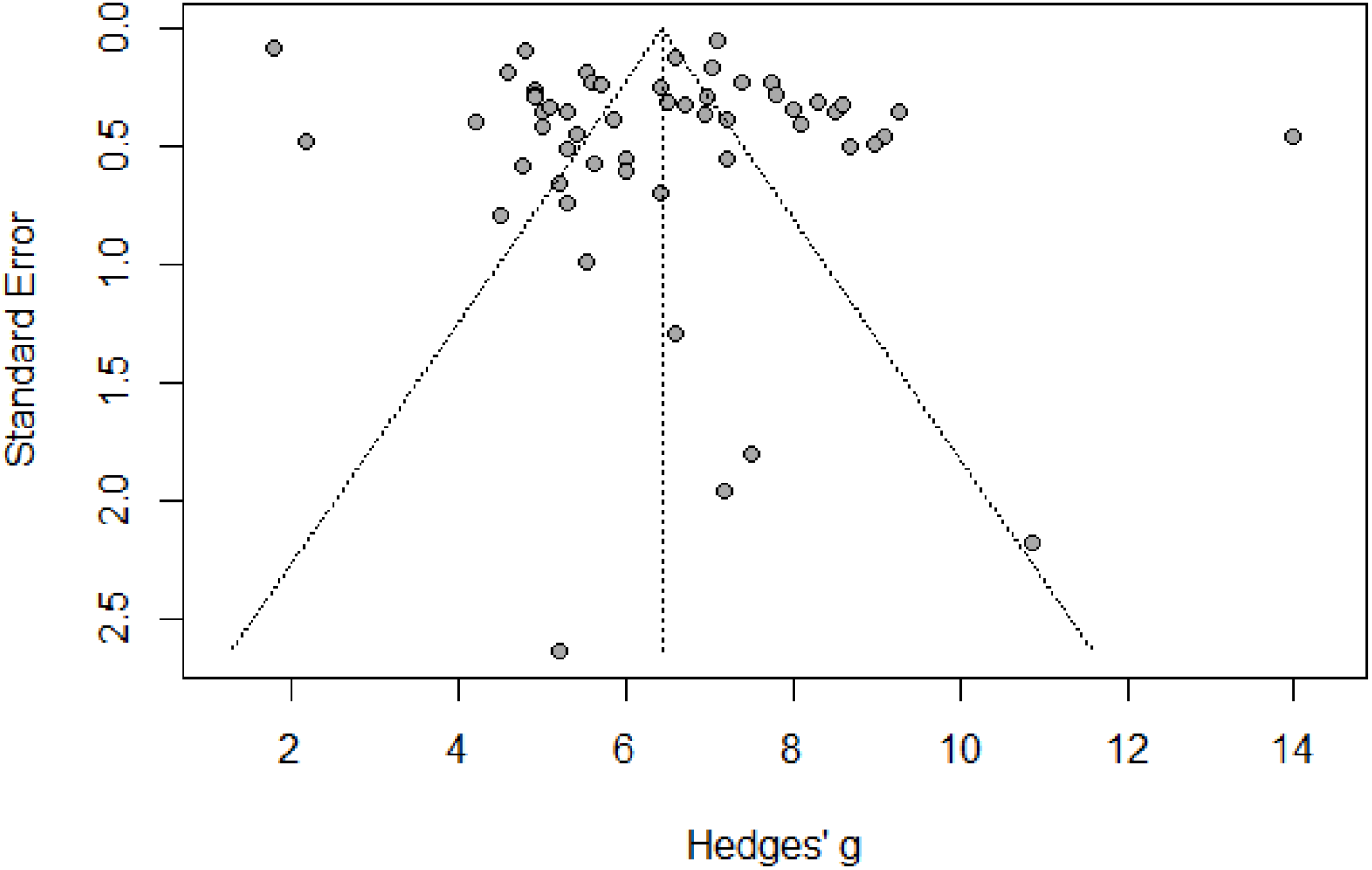
Funnel plot for Analysis 1

### 4.3 Results of Analysis 2

Using the test procedures described in Section 3.2, we assess the 36 transformed results shown in the last 2 columns of Table 4. We obtain that the *p*-value for Cochran’s test is less than 0.01 and *I*^2^ = 95%, suggesting that the random effects model is preferred when conducting meta-analysis. Using the method in Section 3.1 gives us a synthetic approximate mean incubation estimate to be 5.52 days with a 95% CI (5.06, 5.99). Applying the method in Section 3.6 yields the *p*-value 0.43 for the Egger’s test, showing no evidence for the presence of publication bias.

### 4.4 Results of Analysis 3

Combining the 55 studies in *Analysis 1* and 36 studies in *Analysis 2* and applying the test procedures described in Section 3.2 to those 91 studies, we obtain that the *p*-value for Cochran’s test is less than 0.01 and *I*^2^ = 98%, suggesting that the random effects model is preferred when conducting meta-analysis. Figure 7 displays the forest plot of the meta-analysis, showing that the pooled mean incubation estimate for *Analysis 3* is 6.08 days with a 95% CI (5.71, 6.46). By applying the method in Section 3.6, we obtain the *p*-value 0.32 for the Egger’s test, indicating no evidence for the presence of asymmetry in the funnel plot, displayed in Figure 8.

**Figure 7:**
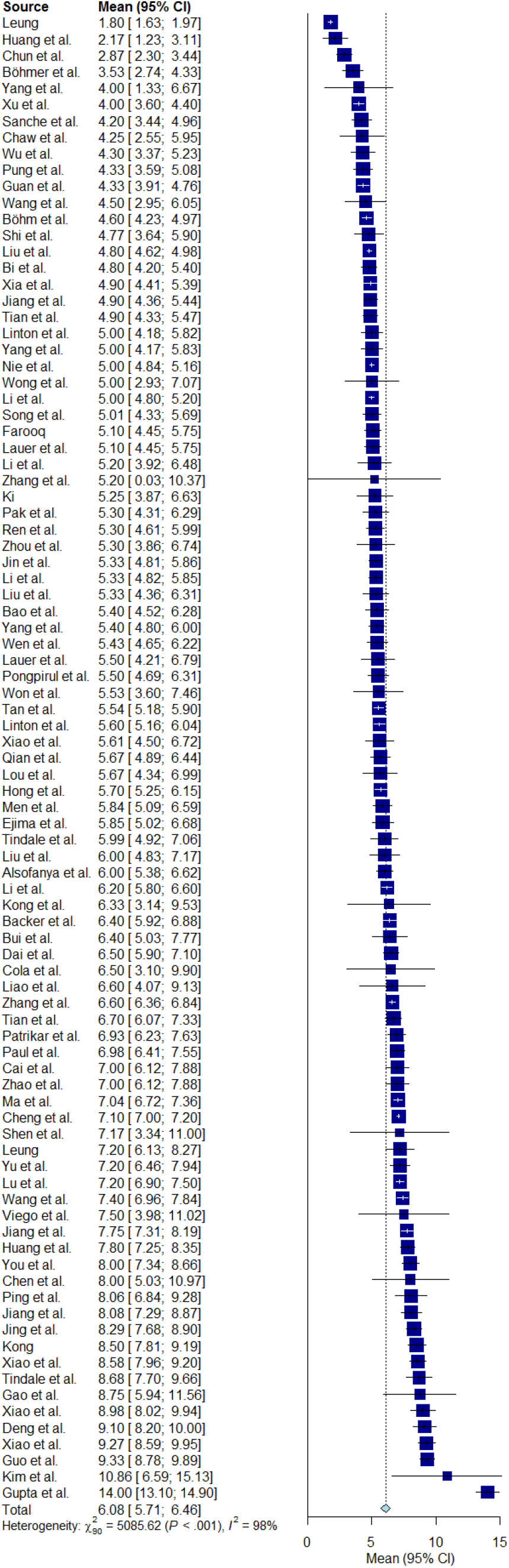
Forest plot for Analysis 3

**Figure 8:**
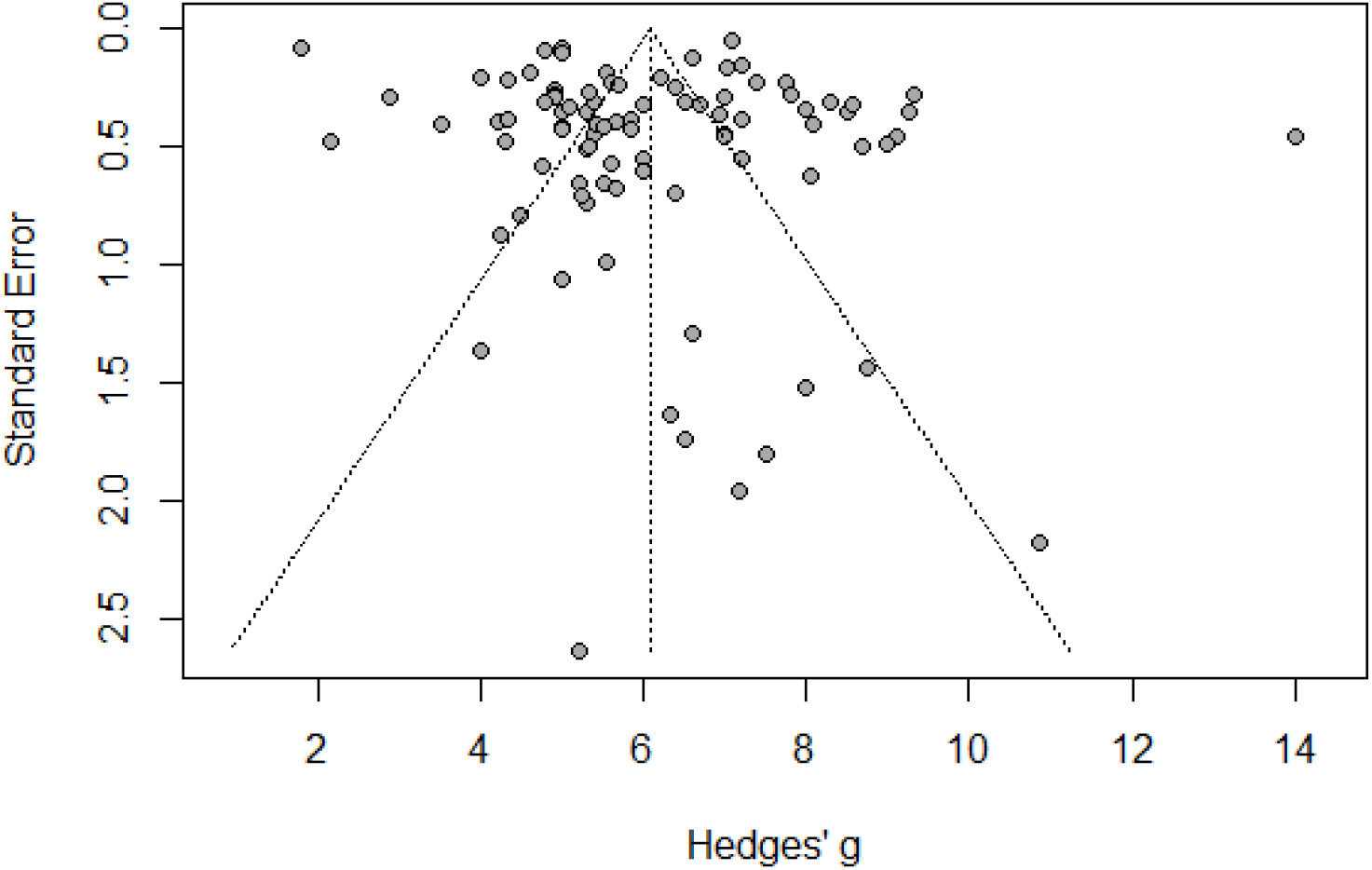
Funnel plot for Analysis 3

### 4.5 Results of Analysis 4

Treating each of the 16 meta-analysis reports in Table 2 as an individual study and using the test procedures described in Section 3.2, we obtain that the *p*-value for Cochran’s test is less than 0.01 and *I*^2^ = 65%, suggesting that the random effects model is preferred when conducting meta-analysis. Using the method in Section 3.1 gives us a synthetic estimate of the mean incubation time for *Analysis 4* to be 5.81 days with a 95% CI (5.57, 6.06). Applying the method in Section 3.6 yields the *p*-value 0.21 for the Egger’s test, suggesting no evidence for the publication bias.

### 4.6 Results of Subgroup Analyses

Applying the test procedures described in Section 3.4 to the 55 studies considered in *Analysis 1*, we further conduct four subgroup analyses, where three grouping strategies are applied based on regions and methodologies as described in Section 2, and another grouping strategy is suggested in Section 4.1 by classifying studies into groups of low, moderate, and high risk of bias. The results are reported in Table 5, where LBCI and UBCI stand for the lower and upper bounds of the 95% confidence intervals related to the mean incubation estimate, respectively.

**Table 5:** Subgroup analysis results

| | $K$ | Mean | LBCI | UBCI | $I^2$ |
| --- | --- | --- | --- | --- | --- |
| *Subgroup Analysis 1* |  |  |  |  |  |
| China | 41 | 6.23 | 5.69 | 6.78 | 99% |
| Mixed1 | 3 | 6.77 | 5.35 | 8.19 | 89% |
| Outside China | 11 | 7.18 | 5.55 | 8.80 | 97% |
| *Subgroup Analysis 2* |  |  |  |  |  |
| Hubei | 9 | 6.01 | 4.55 | 7.47 | 95% |
| Mixed2 | 8 | 5.68 | 4.85 | 6.51 | 97% |
| Outside Hubei | 38 | 6.71 | 6.07 | 7.35 | 97% |
| *Subgroup Analysis 3* |  |  |  |  |  |
| Descriptive Analysis | 14 | 6.22 | 5.12 | 7.33 | 97% |
| Non-parametric | 4 | 8.30 | 4.30 | 12.30 | 99% |
| Parametric | 37 | 6.30 | 5.80 | 6.79 | 99% |
| *Subgroup Analysis 4* |  |  |  |  |  |
| Low Risk | 2 | 5.70 | 4.64 | 6.76 | 15% |
| High Risk | 29 | 6.03 | 5.25 | 6.82 | 99% |
| Moderate Risk | 24 | 6.95 | 6.30 | 7.60 | 95% |

In *Subgroup Analysis 1*, 41 studies are conducted within China, 11 studies are conducted outside China, and 3 studies are based on mixed cases outside and within China (called “Mixed1”). This subgroup analysis suggests a synthetic estimate of the mean incubation time to be 7.18 days (95% CI (5.55, 8.80)) outside China, which is longer than that of within China: 6.23 days with a 95% CI (5.69, 6.78).

For *Subgroup Analysis 2*, 9 studies are conducted within Hubei province in China, 38 studies are conducted outside Hubei province, and 8 studies are conducted based on mixed studies outside and inside Hubei province (called “Mixed2”). The synthetic estimate of the mean incubation time outside Hubei province is 6.71 days (95% CI (6.07, 7.35)), larger than the counterpart inside Hubei, which is 6.01 days (95% CI (4.55, 7.47)).

For *Subgroup Analysis 3*, 14 studies utilized descriptive analysis, 4 studies employed non-parametric models, and 37 assumed utilized parametric models. Among the three methodologies, non-parametric models reveal the largest synthetic estimate as 8.30 days (95% CI (4.30, 12.30)). The rest two methods, descriptive analysis and parametric models, indicate similar estimates of 6.22 days (95% CI (5.12, 7.33)) and 6.30 days (95% CI (5.80, 6.79)), respectively.

Finally, for *Subgroup Analysis 4*, according to Section 4.1, 29 studies are of high risk of bias, 24 studies are of moderate risk, and 2 studies are of low risk of bias. The studies with low risk of bias give a synthetic estimate of 5.70 days (95% CI (4.64, 6.76)), the studies of moderate risk produce an estimate of the pooled mean incubation time to be 6.95 days (95% CI (6.30, 7.60)), and the records from the subgroup of the high risk result in an estimate of 6.03 days (95% CI (5.25, 6.82)).

Further, applying the method of testing the differences between subgroups in Section 3.5, we obtain *p*-values 0.48, 0.15, 0.61, and 0.07 for *Subgroup Analyses 1, 2, 3*, and *4*, respectively, suggesting no significant difference among the subgroups in all four subgroup analyses at the level of 0.05.

### 4.7 Results of Sensitivity Analyses

To further understand the performance of the meta-analysis, we conduct two sensitivity analyses using the same procedure as in *Analysis 1-4*. First, we repeat *Analyses 1* and *3* by adding back those four studies with highly right screwed CIs. Using the methods in Section 3.1, we then respectively obtain synthetic estimates of the mean incubation time to slightly decrease to 6.37 days with a 95% CI (5.86,6.89) and 6.06 days with a 95% CI (5.69, 6.42).

Secondly, we exclude all the studies that have asymmetric CIs in *Analysis 1* and conduct another meta-analysis on the 13 remaining studies. The resultant synthetic estimate of the time is 6.06 days with 95% CI (5.27,6.85), which is smaller than that of *Analysis 1* and almost identical to the estimate from *Analysis 3* with a wider CI.

## 5 Conclusions and Discussion

To provide a sensible understanding of the average incubation time for COVID-19, in this article we take different angles to examine the reported estimates in the literature that were obtained from different studies. With the 55 estimates of the mean incubation time of COVID-19, we obtain that the synthetic estimate from meta-analysis is 6.43 days (95% CI (5.90, 6.96)). Further combined with 36 estimates transformed from the reported estimates of the median incubation time of COVID-19, the meta-analysis yields a synthetic estimate of the mean incubation time decreased to 6.08 days (95% CI (5.71, 6.46)).

Subgroup analyses suggest that the estimate of the mean incubation time is 7.18 days (95% CI (5.55, 8.80)) and 6.71 days (95% CI (6.07, 7.35)), respectively, for patients outside China and outside Hubei province. For different risk levels, studies with low risk of bias yield the smallest pooled mean estimate of 5.70 days (95% CI (4.64, 6.76)), compared to moderate and high risk groups. Notably, none of the between-subgroup differences is significant. Moreover, the largest pooled average result is obtained from non-parametric models, which is 8.30 days (95% CI (4.30, 12.30)).

Sensitivity analyses suggest that if including or excluding studies with highly skewed CIs considerably changes estimates. While it is difficult to determine exactly the average incubation time of COVID-19, our study here provides insights into understating of this unknown quantity by incorporating various features of the available estimates, including heterogeneity, varying sample sizes, study bias, differences in estimation methods, and so on.

This being said, there are limitations in the analyses here, just like other available studies. Although our search of the literature spans the period of between January 1, 2020, and May 20, 2021, the reported estimates of the mean incubation time of COVID-19 are mainly obtained from the studies of those infected cases prior to March 31, 2020. The results thereby do not reflect the feature that the average incubation time may change with the emerging variants of the virus. The normality assumption in the meta-analysis may not be valid since the distribution of incubation times is assumed to be right-skewed in some analyses. Many studies did not give individual characteristics such as age, the sex ratio, and medical conditions of patients, which hinders us from further exploring the heterogeneity of the studies. Most studies using parametric models assumed a distribution such as Gamma, Weibull, or log-normal to describe COVID-19 incubation times. Such distributional assumptions, however, are basically not testable.

## Supporting information

Supplemental Whole

## Data Availability

All data produced in the present work are contained in the manuscript.

## Acknowledgements

The research was partially supported by the grants of the Discovery Grants Program and the Emerging Infectious Disease Modeling Program from the Natural Sciences and Engineering Research Council of Canada. Yi is Canada Research Chair in Data Science (Tier 1). Her research was undertaken, in part, thanks to funding from the Canada Research Chairs program.

## Authors’ Contributions

YW searches the data, conducts the analysis, and prepares an initial draft. Professor GYY offers ideas for the project and writes the manuscript. All authors have read and approved the manuscript.

## Ethical Approval

Not required.

