## Supplemental Whole for "Estimation of the COVID-19 Average Incubation Time: Systematic Review, Meta-analysis and Sensitivity Analyses"

#### S1: Risk of Bias Checklist

Figures S.1-S.3 present the risk of bias checklists which adapt the checklist of Hoy et al. (2012), where we modify their items 9 and 10 about disease prevalence to describe COVID-19 incubation times as suggested by Quesada et al. (2021).

Figure S.1: Checklist for risk of bias assessment-1

This tool is designed to assess the risk of bias in population-based prevalence studies. Please read the additional notes for each item when initially using the tool. Note: If there is insufficient information in the article to permit a judgment for a particular item, please answer **No (HIGH RISK)** for that particular item.

| Risk of bias item | Criteria for answers (please circle one option) | Additional notes and examples |
|---|---|---|
| ***External Validity*** | | |
| 1. Was the study's target population **a close representation** of the national population in relation to relevant variables, e.g. age, sex, occupation? | • **Yes (LOW RISK):** The study's target population was a close representation of the national population.<br>• **No (HIGH RISK)**: The study's target population was clearly NOT representative of the national population. | The **target population** refers to the group of people or entities to which the results of the study will be generalized. Examples:<br>• The study was a national health survey of people 15 years and over and the sample was drawn from a list that included all individuals in the population aged 15 years and over. The answer is: **Yes (LOW RISK).**<br>• The study was conducted in one province only, and it is not clear if this was representative of the national population. The answer is: **No (HIGH RISK).**<br>• The study was undertaken in one village only and it is clear this was not representative of the national population. The answer is: **No (HIGH RISK).** |
| 2. Was the sampling frame a **true or close representation** of the target population? | • **Yes (LOW RISK):** The sampling frame was a **true or close** representation of the target population.<br>• **No (HIGH RISK)**: The sampling frame was NOT a **true or close** representation of the target population. | The **sampling frame** is a list of the sampling units in the target population and the study sample is drawn from this list. Examples:<br>• The sampling frame was a list of almost every individual within the target population. The answer is: **Yes (LOW RISK)**.<br>• The cluster sampling method was used and the sample of clusters/villages was drawn from a list of all villages in the target population. The answer is: **Yes (LOW RISK)**.<br>• The sampling frame was a list of just one particular ethnic group within the overall target population, which comprised many groups. The answer is: **No (HIGH RISK).** |
| 3. Was some form of **random selection** used to select the sample, OR, was a census undertaken? | • **Yes (LOW RISK):** A census was undertaken, OR, some form of random selection was used to select the sample (e.g. simple random sampling, stratified random sampling, cluster sampling, systematic sampling).<br>• **No (HIGH RISK)**: A census was NOT undertaken, AND some form of random selection was NOT used to select the sample. | A census collects information from every unit in the sampling frame. In a survey, only part of the sampling frame is sampled. In these instances, random selection of the sample helps minimize study bias. Examples:<br>• The sample was selected using simple random sampling. The answer is: **Yes (LOW RISK)**.<br>• The target population was the village and every person in the village was sampled. The answer is: **Yes (LOW RISK)**.<br>• The nearest villages to the capital city were selected in order to save on the cost of fuel. The answer is: **No (HIGH RISK).** |
| 4. Was the likelihood of **non-response bias minimal?** | • **Yes (LOW RISK):** The response rate for the study was >/=75%, OR, an analysis was performed that showed no significant difference in relevant demographic characteristics between responders and non-responders<br>• **No (HIGH RISK)**: The response rate was <75%, and if any analysis comparing responders and non-responders was done, it showed a significant difference in relevant demographic characteristics between responders and non-responders. | Examples:<br>• The response rate was 68%; however, the researchers did an analysis and found no significant difference between responders and non-responders in terms of age, sex, occupation and socio-economic status. The answer is: **Yes (LOW RISK)**.<br>• The response rate was 65% and the researchers did NOT carry out an analysis to compare relevant demographic characteristics between responders and non-responders. The answer is: **No (HIGH RISK)**.<br>• The response rate was 69% and the researchers did an analysis and found a significant difference in age, sex and socio-economic status between responders and non-responders. The answer is: **No (HIGH RISK)**. |

Figure S.2: Checklist for risk of bias assessment-2

| ***Internal Validity*** | | |
|---|---|---|
| 5. Were data collected **<u>directly from the subjects</u>** (as opposed to a proxy)? | • **Yes (LOW RISK):** All data were collected directly from the subjects.<br>• **No (HIGH RISK)**: In some instances, data were collected from a proxy. | A proxy is a representative of the subject. Examples:<br>• All eligible subjects in the household were interviewed separately. The answer is: **Yes (LOW RISK)**.<br>• A representative of the household was interviewed and questioned about the presence of low back pain in each household member. The answer is: **No (HIGH RISK)**. |
| 6. Was an **<u>acceptable case definition</u>** used in the study? | • **Yes (LOW RISK):** An acceptable case definition was used.<br>• **No (HIGH RISK)**: An acceptable case definition was <u>NOT</u> used. | • For a study on low back pain, the following case definition was used: "Low back pain is defined as activity-limiting pain lasting more than one day in the area on the posterior aspect of the body from the bottom of the 12th rib to the lower gluteal folds." The answer is: **Yes (LOW RISK)**.<br>• For a study on back pain, there was no description of the specific anatomical location 'back' referred to. The answer is: **No (HIGH RISK)**.<br>• For a study on osteoarthritis, the following case definition was used: "Symptomatic osteoarthritis of the hip or knee, radiologically confirmed as Kellgren-Lawrence grade 2-4". The answer is: LOW RISK. |
| 7. Was the study instrument that measured the parameter of interest (e.g. prevalence of low back pain) shown to have **<u>reliability and validity (if necessary)</u>**? | • **Yes (LOW RISK):** The study instrument had been shown to have reliability and validity (if this was necessary), e.g. test-re-test, piloting, validation in a previous study, etc.<br>• **No (HIGH RISK)**: The study instrument had <u>NOT</u> been shown to have reliability or validity (if this was necessary). | • The authors used the COPCORD questionnaire, which had previously been validated. They also tested the inter-rater reliability of the questionnaire. The answer is: **Yes (LOW RISK)**.<br>• The authors developed their own questionnaire and did not test this for validity or reliability. The answer is: **No (HIGH RISK)**. |
| 8. Was the **<u>same mode of data collection</u>** used for all subjects? | • **Yes (LOW RISK):** The same mode of data collection was used for all subjects.<br>• **No (HIGH RISK)**: The same mode of data collection was NOT used for all subjects. | The mode of data collection is the method used for collecting information from the subjects. The most common modes are face-to-face interviews, telephone interviews and self-administered questionnaires. Examples:<br>• All eligible subjects had a face-to-face interview. The answer is: **Yes (LOW RISK)**.<br>• Some subjects were interviewed over the telephone and some filled in postal questionnaires. The answer is: **No (HIGH RISK)**. |
| 9. Was the **<u>start and end of incubation time</u>** clearly identified? | • **Yes (LOW RISK):** The start and end of incubation time were clearly identified in the reports.<br>• **No (HIGH RISK)**: The start or end of incubation time was not clearly identified in the reports. | The estimation of incubation time (days) requires the date of exposure to SARS-CoV-2 and the date of symptom onset. Sometimes data were censored and no clear dates can be obtained. Examples:<br>• The start and end of the incubation time for all the subjects are not censored and were clearly identified. The answer is: **Yes (LOW RISK)**.<br>• The start and end of the incubation time for some the subjects were censored. The answer is: **No (HIGH RISK)**.<br>• The start and end of the incubation time were not censored, but the exact dates were not clearly identified in the reports. The answer is: **No (HIGH RISK)**. |
| 10. Was the **<u>exposure</u>** of the patient clearly identified? | • **Yes (LOW RISK):** The paper presented exposures that were clearly identified.<br>• **No (HIGH RISK)**: The paper included some cases for which the exposures were not clearly identified. | The exposure of an individual refers to a food vehicle or a patient source. Examples:<br>• The reports included the identification of implicated Food vehicle or source patient for each individual. The answer is: **Yes (LOW RISK)**.<br>• There exist some cases that did not have any information about the food vehicle or source patient. The answer is: **No (HIGH RISK).** |
| ***11. Summary item on the overall risk of study bias*** | | |
| • **LOW RISK OF BIAS:** Further research is <u>very unlikely</u> to change our confidence in the estimate.<br>• **MODERATE RISK OF BIAS:** Further research is <u>likely</u> to have an important impact on our confidence in the estimate and may change the estimate. | | |

Figure S.3: Checklist for risk of bias assessment-3

- **HIGH RISK OF BIAS:** Further research is very likely to have an important impact on our confidence in the estimate and is likely to change the estimate.

### S2: Transformation Formulas for Median Estimates

To obtain an estimate of the mean incubation time and associated standard deviation (SD) from reported estimates related to the median incubation time, we apply the approximation method discussed by Wan et al. (2014).

To be specific, let $n$ denote the sample size of a study and let $\Phi(\cdot)$ denote the cumulative distribution function of the standard normal distribution. If the study reports estimates of the median, first quartile and third quartile, denoted $Q_2$, $Q_1$ and $Q_3$, respectively, then estimates of the mean and SD may be approximated by:

$$\widehat{mean} = \frac{Q_1 + Q_2 + Q_3}{3} \text{ and } \widehat{SD} = \frac{Q_3 - Q_1}{2\Phi^{-1}(\frac{0.75n-0.125}{n+0.25})}. \tag{1}$$

If the study reports estimates of the median, the lower and upper bounds of the range, denoted $Q_2$, $Q_{min}$ and $Q_{max}$, respectively, then estimates of the mean and SD can be given by:

$$\widehat{mean} = \frac{Q_{min} + 2Q_2 + Q_{max}}{4} \text{ and } \widehat{SD} = \frac{Q_{max} - Q_{min}}{2\Phi^{-1}(\frac{n-0.375}{n+0.25})}. \tag{2}$$

### S3: Information of the Collected Papers

Alene, M., Yismaw, L., Assemie, M. A., Ketema, D. B., Gietaneh, W., and Birhan, T. Y. (2021). Serial interval and incubation period of COVID-19: A systematic review and meta-analysis. *BMC Infectious Diseases*, 21(1):1–9.

Alsofayan, Y. M., Althunayyan, S. M., Khan, A. A., Hakawi, A. M., and Assiri, A. M. (2020). Clinical characteristics of COVID-19 in saudi arabia: A national retrospective study. *Journal of Infection and Public Health*, 13(7):920–925.

Backer, J. A., Klinkenberg, D., and Wallinga, J. (2020). Incubation period of 2019 novel coronavirus (2019-ncov) infections among travellers from Wuhan, China, 20–28 January 2020. *Eurosurveillance*, 25(5):2000062.

Banka, P. and Comiskey, C. (2020). The incubation period of COVID-19: A scoping review and meta-analysis to aid modelling and planning. *medRxiv*.

Bao, C., Pan, E., Ai, J., Dai, Q., Xu, K., Shi, N., Gao, Q., Hu, J., Peng, Z., Huang, H., Jin, H., and Zhu, F. (2021). COVID-19 outbreak following a single patient exposure at an entertainment site: An epidemiological study. *Transboundary and Emerging Diseases*, 68(2):773–781.

Bi, Q., Wu, Y., Mei, S., Ye, C., Zou, X., Zhang, Z., Liu, X., Wei, L., Truelove, S. A., Zhang, T., Gao, W., Cheng, C., Tang, X., Wu, X., Wu, Y., Sun, B., Huang, S., Sun, Y., Zhang, J., Ma, T., Lessler, J., and Feng, T. (2020). Epidemiology and transmission of COVID-19 in 391 cases and 1286 of their close contacts in Shenzhen, China: a retrospective cohort study. *The Lancet Infectious Diseases*, 20(8):911–919.

Bui, L. V., Nguyen, H. T., Levine, H., Nguyen, H. N., Nguyen, T.-A., Nguyen, T. P., Nguyen, T. T., Do, T. T. T., Pham, N. T., and Bui, M. H. (2020). Estimation of the incubation period of COVID-19 in vietnam. *PLOS ONE*, 15(12):1–9.

Böhm, S., Woudenberg, T., Chen, D., Marosevic, D. V., Böhmer, M. M., Hansen, L., Wallinga, J., Sing, A., and Katz, K. (2021). Epidemiology and transmission characteristics of early COVID-19 cases, 20 January–19 March 2020, in Bavaria, Germany. *Epidemiology and Infection*, 149:e65.

Böhmer, M. M., Buchholz, U., Corman, V. M., Hoch, M., Katz, K., Marosevic, D. V., Böhm, S., Woudenberg, T., Ackermann, N., Konrad, R., Eberle, U., Treis, B., Dangel, A., Bengs, K., Fingerle, V., Berger, A., Hörmansdorfer, S., Ippisch, S., Wicklein, B., Grahl, A., Pörtner, K., Muller, N., Zeitlmann, N., Boender, T. S., Cai, W., Reich, A., an der Heiden, M., Rexroth, U., Hamouda, O., Schneider, J., Veith, T., Mühlemann, B., Wölfel,

R., Antwerpen, M., Walter, M., Protzer, U., Liebl, B., Haas, W., Sing, A., Drosten, C., and Zapf, A. (2020). Investigation of a COVID-19 outbreak in germany resulting from a single travel-associated primary case: A case series. *The Lancet Infectious Diseases*, 20(8):920–928.

Cai, Y., Liu, J., Yang, H., Wang, M., Guo, Q., Huang, D., Yu, Q., and Xiao, E. (2020). Association between incubation period and clinical characteristics of patients with COVID-19. *Journal of International Medical Research*, 48(9):0300060520956834.

Chaw, L., Koh, W. C., Jamaludin, S. A., Naing, L., Alikhan, M. F., and Wong, J. (2020). Analysis of SARS-CoV-2 transmission in different settings, brunei. *Emerging infectious diseases*, 26(11):2598.

Chen, G., Wu, M., Qin, C., Wu, B., Luo, W., Liu, L., and Liu, J. (2020). Epidemiological analysis of 18 patients with COVID-19. *European Review for Medical and Pharmacological Sciences*, 24(23):12522–12526.

Cheng, C., Zhang, D., Dang, D., Geng, J., Zhu, P., Yuan, M., Liang, R., Yang, H., Jin, Y., Xie, J., Chen, S., and Duan, G. (2021). Incubation period of COVID-19 from 11545 patients in observation study. Available at Research Square: https://europepmc.org/article/PPR/PPR307228.

Cheng, H., Jian, S., Liu, D., Ng, T.-C., Huang, W., Lin, H.-H., and for the Taiwan COVID-19 Outbreak Investigation Team (2020). Contact tracing assessment of COVID-19 transmission dynamics in taiwan and risk at different exposure periods before and after symptom onset. *JAMA Internal Medicine*, 180(9):1156–1163.

Chun, J. Y., Baek, G., and Kim, Y. (2020). Transmission onset distribution of COVID-19. *International Journal of Infectious Diseases*, 99:403–407.

Dai, J., Yang, L., and Zhao, J. (2020). Probable longer incubation period for elderly COVID-

19 cases: analysis of 180 contact tracing data in Hubei province, China. *Risk Management and Healthcare Policy*, 13:1111–1117.

Deng, Y., You, C., Liu, Y., Qin, J., and Zhou, X.-H. (2020). Estimation of incubation period and generation time based on observed length-biased epidemic cohort with censoring for COVID-19 outbreak in China. *Biometrics*, 77(3):929–941.

Dhouib, W., Maatoug, J., Ayouni, I., Zammit, N., Ghammem, R., Fredj, S. B., and Ghannem, H. (2021). The incubation period during the pandemic of COVID-19: A systematic review and meta-analysis. *Systematic reviews*, 10(1):1–14.

Du, Z., Gu, J., Li, J., Lin, X., Wang, Y., Chen, L., and Hao, Y. (2020). Estimating the distribution of covid-19 incubation period by interval-censored data estimation method. *Chinese Journal of Epidemiology*, 41(7):1000—1003.

Ejima, K., Kim, K. S., Ludema, C., Bento, A. I., Iwanami, S., Fujita, Y., Ohashi, H., Koizumi, Y., Watashi, K., Aihara, K., Nishiura, H., and Iwami, S. (2021). Estimation of the incubation period of COVID-19 using viral load data. *Epidemics*, 35:100454.

Elias, C., Sekri, A., Leblanc, P., Cucherat, M., and Vanhems, P. (2021). The incubation period of COVID-19: A meta-analysis. *International Journal of Infectious Diseases*, 104:708–710.

Farooq, M. H. (2021). The incubation period of coronavirus disease 2019 from publicly reported confirmed cases: Estimation and application. Available at: https://studentspectrum.imdcollege.edu.pk/2021/(14)%20The%20Incubation%20period%20(M.%20Hassan%20Farooq)%20-%20Copy.pdf.

Gao, Y., Shi, C., Chen, Y., Shi, P., Liu, J., Xiao, Y., Shen, Y., and Chen, E. (2020). A cluster of the corona virus disease 2019 caused by incubation period transmission in Wuxi, China. *Journal of Infection*, 80(6):666–670.

Guan, W., Ni, Z., Hu, Y., Liang, W., Ou, C., He, J., Liu, L., Shan, H., Lei, C., Hui, D. S., Du, B., Li, L., Zeng, G., Yuen, K., Chen, R., Tang, C., Wang, T., Chen, P., Xiang, J., Li, S., Wang, J., Liang, Z., Peng, Y., Wei, L., Liu, Y., Hu, Y., Peng, P., Wang, J., Liu, J., Chen, Z., Li, G., Zheng, Z., Qiu, S., Luo, J., Ye, C., Zhu, S., and Zhong, N. (2020). Clinical characteristics of coronavirus disease 2019 in China. *New England Journal of Medicine*, 382(18):1708–1720.

Guo, C., He, L., Yin, J., Meng, X., Tan, W., Yang, G., Bo, T., Liu, J., Lin, X., and Chen, X. (2020). Epidemiological and clinical features of pediatric COVID-19. *BMC Medicine*, 18(1):1–7.

Gupta, A., Pradhan, B., and Maulud, K. N. A. (2020). Estimating the impact of daily weather on the temporal pattern of COVID-19 outbreak in india. *Earth Systems and Environment*, 4(3):523–534.

He, W., Yi, G. Y., and Zhu, Y. (2020). Estimation of the basic reproduction number, average incubation time, asymptomatic infection rate, and case fatality rate for COVID-19: Meta-analysis and sensitivity analysis. *Journal of Medical Virology*, 92(11):2543–2550.

Hong, H., Shi, H., Jiang, H., Gu, X., Chen, Y., Ding, K., and Xu, G. (2020). Epidemic dynamic model based evaluation of effectiveness of prevention and control strategies for COVID-19 in Ningbo. *Chinese Epidemiology Journal*, 41(10):1606–1610.

Huang, L., Zhang, X., Zhang, X., Wei, Z., Zhang, L., Xu, J., Liang, P., Xu, Y., Zhang, C., and Xu, A. (2020). Rapid asymptomatic transmission of COVID-19 during the incubation period demonstrating strong infectivity in a cluster of youngsters aged 16-23 years outside Wuhan and characteristics of young patients with COVID-19: A prospective contact-tracing study. *Journal of Infection*, 80(6):e1–e13.

Huang, S., Li, J., Dai, C., Tie, Z., Xu, J., Xiong, X., Hao, X., Wang, Z., and Lu, C.

(2021). Incubation period of coronavirus disease 2019: new implications for intervention and control. *International Journal of Environmental Health Research*, 0(0):1–9.

Jiang, A. B.-Z., Lieu, R., and Quenby, S. (2020a). Significantly longer COVID-19 incubation times for the elderly, from a case study of 136 patients throughout China. *medRxiv*.

Jiang, X., Rayner, S., and Luo, M.-H. (2020b). Does SARS-CoV-2 has a longer incubation period than SARS and MERS? *Journal of Medical Virology*, 92(5):476–478.

Jiang, Z., Yang, B., Qin, J., and Zhou, Y. (2021). Enhanced empirical likelihood estimation of incubation period of COVID-19 by integrating published information. *Statistics in Medicine*, 40(19):4252–4268.

Jin, X., Lian, J.-S., Hu, J., Gao, J., Zheng, L., Zhang, Y., Hao, S., Jia, H., Cai, H., Zhang, X., Yu, G., Xu, K., Wang, X., Gu, J., Zhang, S., Ye, C., Jin, C., Lu, Y., Yu, X., Yu, X., Huang, J., Xu, K., Ni, Q., Yu, C., Zhu, B., Li, Y., Liu, J., Zhao, H., Zhang, X., Yu, L., Guo, Y., Su, J., Tao, J., Lang, G., Wu, X., Wu, W., Qv, T., Xiang, D., Yi, P., Shi, D., Chen, Y., Ren, Y., Qiu, Y., Li, L., Sheng, J., and Yang, Y. (2020). Epidemiological, clinical and virological characteristics of 74 cases of coronavirus-infected disease 2019 (COVID-19) with gastrointestinal symptoms. *Gut*, 69(6):1002–1009.

Khalili, M., Karamouzian, M., Nasiri, N., Javadi, S., Mirzazadeh, A., and Sharifi, H. (2020). Epidemiological characteristics of COVID-19: a systematic review and meta-analysis. *Epidemiology and Infection*, 148:e130.

Ki, M. and for 2019-nCoV, T. F. (2020). Epidemiologic characteristics of early cases with 2019 novel coronavirus (2019-nCoV) disease in Korea. *Epidemiology and health*, 42:e2020007.

Kim, S. E., Jeong, H. S., Yu, Y., Shin, S. U., Kim, S., Oh, T. H., Kim, U. J., Kang, S., Jang, H., Jung, S., and Park, K. (2020). Viral kinetics of SARS-CoV-2 in asymptomatic carriers and presymptomatic patients. *International Journal of Infectious Diseases*, 95:441–443.

Kong, D., Zheng, Y., Wu, H., Pan, H., Wagner, A. L., Zheng, Y., Gong, X., Zhu, Y., Jin, B., Xiao, W., Mao, S., Lin, S., Han, R., Yu, X., Cui, P., Jiang, C., Fang, Q., Lu, Y., and Fu, C. (2020). Pre-symptomatic transmission of novel coronavirus in community settings. *Influenza and Other Respiratory Viruses*, 14(6):610–614.

Kong, T.-k. (2020). Longer incubation period of coronavirus disease 2019 (COVID-19) in older adults. *Aging Medicine*, 3(2):102–109.

Lauer, S. A., Grantz, K. H., Bi, Q., Jones, F. K., Zheng, Q., Meredith, H. R., Azman, A. S., Reich, N. G., and Lessler, J. (2020). The incubation period of coronavirus disease 2019 (COVID-19) from publicly reported confirmed cases: Estimation and application. *Annals of Internal Medicine*, 172(9):577–582.

Lee, H., Kim, K., Choi, K., Hong, S., Son, H., and Ryu, S. (2020). Incubation period of the coronavirus disease 2019 (COVID-19) in busan, south korea. *Journal of Infection and Chemotherapy*, 26(9):1011–1013.

Leung, C. (2020). The difference in the incubation period of 2019 novel coronavirus (SARS-CoV-2) infection between travelers to Hubei and nontravelers: The need for a longer quarantine period. *Infection Control & Hospital Epidemiology*, 41(5):594–596.

Li, J., Ding, J., Chen, L., Hong, L., Yu, X., Ye, E., Sun, G., Zhang, B., Zhang, X., and Sun, Q. (2020a). Epidemiological and clinical characteristics of three family clusters of COVID-19 transmitted by latent patients in China. *Epidemiology and Infection*, 148:e137.

Li, J., Huang, D. Q., Zou, B., Yang, H., Hui, W. Z., Rui, F., Yee, N. T. S., Liu, C., Nerurkar, S. N., Kai, J. C. Y., Teng, M. L. P., Li, X., Zeng, H., Borghi, J. A., Henry, L., Cheung, R., and Nguyen, M. H. (2021a). Epidemiology of COVID-19: A systematic review and meta-analysis of clinical characteristics, risk factors, and outcomes. *Journal of Medical Virology*, 93(3):1449–1458.

Li, M., Chen, P., Yuan, Q., Song, B., and Ma, J. (2020b). Transmission characteristics of the COVID-19 outbreak in China: A study driven by data. *medRxiv*.

Li, Q., Guan, X., Wu, P., Wang, X., Zhou, L., Tong, Y., Ren, R., Leung, K. S., Lau, E. H., Wong, J. Y., Xing, X., Xiang, N., Wu, Y., Li, C., Chen, Q., Li, D., Liu, T., Zhao, J., Liu, M., Tu, W., Chen, C., Jin, L., Yang, R., Wang, Q., Zhou, S., Wang, R., Liu, H., Luo, Y., Liu, Y., Shao, G., Li, H., Tao, Z., Yang, Y., Deng, Z., Liu, B., Ma, Z., Zhang, Y., Shi, G., Lam, T. T., Wu, J. T., Gao, G. F., Cowling, B. J., Yang, B., Leung, G. M., and Feng, Z. (2020c). Early transmission dynamics in Wuhan, China, of novel coronavirus–infected pneumonia. *New England Journal of Medicine*, 382(13):1199–1207.

Li, Z., Zhang, Y., Peng, L., Gao, R., Jing, J., Wang, J., Ren, B., Xu, J., and Wang, T. (2021b). Demand for longer quarantine period among common and uncommon COVID-19 infections: a scoping review. *Infectious Diseases of Poverty*, 10(1):1–9.

Liao, J., Fan, S., Chen, J., Wu, J., Xu, S., Guo, Y., Li, C., Zhang, X., Wu, C., Mou, H., Song, C., Li, F., Wu, G., Zhang, J., Guo, L., Liu, H., Lv, J., Xu, L., and Lang, C. (2020). Epidemiological and clinical characteristics of COVID-19 in adolescents and young adults. *The Innovation*, 1(1):100001.

Linton, N. M., Kobayashi, T., Yang, Y., Hayashi, K., Akhmetzhanov, A. R., Jung, S.-m., Yuan, B., Kinoshita, R., and Nishiura, H. (2020). Incubation period and other epidemiological characteristics of 2019 novel coronavirus infections with right truncation: A statistical analysis of publicly available case data. *Journal of Clinical Medicine*, 9(2):538.

Liu, J., Liao, X., Qian, S., Yuan, J., Wang, F., Liu, Y., Wang, Z., Wang, F., Liu, L., and Zhang, Z. (2020a). Community transmission of severe acute respiratory syndrome coronavirus 2, Shenzhen, China, 2020. *Emerging Infectious Diseases*, 26(6):1320.

Liu, J.-Y., Chen, T.-J., and Hwang, S.-J. (2020b). Analysis of community-acquired COVID-19 cases in taiwan. *Journal of the Chinese Medical Association*, 83(12):1087–1092.

Liu, T., Hu, J., Kang, M., Lin, L., Zhong, H., Xiao, J., He, G., Song, T., Huang, Q., Rong, Z., et al. (2020c). Transmission dynamics of 2019 novel coronavirus (2019-ncov). Avaliable at SSRN: https://ssrn.com/abstract=3526307.

Lou, B., Li, T.-D., Zheng, S., Su, Y., Li, Z., Liu, W., Yu, F., Ge, S., Zou, Q., Yuan, Q., Lin, S., Hong, C., Yao, X., Zhang, X., Wu, D., Zhou, G., Hou, W., Li, T., Zhang, Y., Zhang, S., Fan, J., Zhang, J., Xia, N.-S., and Chen, Y. (2020a). Epidemiological parameters of COVID-19 and its implication for infectivity among patients in China, 1 January to 11 February 2020. *Eurosurveillance*, 25(40):2000250.

Lou, B., Li, T.-D., Zheng, S.-F., Su, Y.-Y., Li, Z.-Y., Liu, W., Yu, F., Ge, S.-X., Zou, Q.-D., Yuan, Q., Lin, S., Hong, C.-M., Yao, X.-Y., Zhang, X.-J., Wu, D.-H., Zhou, G.-L., Hou, W.-H., Li, T.-T., Zhang, Y.-L., Zhang, S.-Y., Fan, J., Zhang, J., Xia, N.-S., and Chen, Y. (2020b). Serology characteristics of SARS-CoV-2 infection after exposure and post-symptom onset. *European Respiratory Journal*, 56(2):0903–1936.

Ma, S., Zhang, J., Zeng, M., Yun, Q., Guo, W., Zheng, Y., Zhao, S., Wang, M. H., and Yang, Z. (2020). Epidemiological parameters of COVID-19: Case series study. *Journal of Medical Internet Research*, 22(10):e19994.

McAloon, C., Collins, Á., Hunt, K., Barber, A., Byrne, A. W., Butler, F., Casey, M., Griffin, J., Lane, E., McEvoy, D., Wall, P., Green, M., O'Grady, L., and More, S. J. (2020). Incubation period of COVID-19: A rapid systematic review and meta-analysis of observational research. *BMJ Open*, 10(8):e039652.

Men, K., Wang, X., Yihao, L., Zhang, G., Hu, J., Gao, Y., and Han, H. (2020). Estimate the incubation period of coronavirus 2019 (COVID-19). *medRxiv*.

Nie, X., Fan, L., Mu, G., Tan, Q., Wang, M., Xie, Y., Cao, L., Zhou, M., Zhang, Z., and Chen, W. (2020). Epidemiological characteristics and incubation period of 7015 confirmed cases with coronavirus disease 2019 outside Hubei province in China. *The Journal of Infectious Diseases*, 222(1):26–33.

Ondee, T., Pongpirul, K., Visitchanakun, P., Saisorn, W., Kanacharoen, S., Wongsaroj, L., Kullapanich, C., Ngamwongsatit, N., Settachaimongkon, S., Somboonna, N., et al. (2021). Lactobacillus acidophilus la5 improves saturated fat-induced obesity mouse model through the enhanced intestinal akkermansia muciniphila. *Scientific Reports*, 11(1):1–16.

Pak, D., Langohr, K., Ning, J., Cortés Martínez, J., Gómez Melis, G., and Shen, Y. (2020). Modeling the coronavirus disease 2019 incubation period: Impact on quarantine policy. *Mathematics*, 8(9):1631.

Patrikar, S., Kotwal, A., Bhatti, V., Banerjee, A., Chatterjee, K., Kunte, R., and Tambe, M. (2020). Incubation period and reproduction number for novel coronavirus (COVID-19) infections in india. *medRxiv*.

Paul, S. and Lorin, E. (2020). Distribution of incubation period of COVID-19 in the canadian context: Modeling and computational study. *medRxiv*.

Ping, K., Lei, M., Gou, Y., Tao, Y., and Huang, Y. (2020). Epidemiologic characteristics of COVID-19 in Guizhou, China. *medRxiv*.

Pormohammad, A., Ghorbani, S., Khatami, A., Razizadeh, M. H., Alborzi, E., Zarei, M., Idrovo, J.-P., and Turner, R. J. (2021). Comparison of influenza type a and b with COVID-19: A global systematic review and meta-analysis on clinical, laboratory and radiographic findings. *Reviews in Medical Virology*, 31(3):e2179.

Pung, R., Chiew, C. J., Young, B. E., Chin, S., Chen, M. I.-C., Clapham, H. E., Cook, A. R., Maurer-Stroh, S., Toh, M. P. H. S., Poh, C., Low, M., Lum, J., Koh, V. T. J., Mak, T. M., Cui, L., Lin, R. V. T. P., Heng, D., Leo, Y.-S., Lye, D. C., Lee, V. J. M., qian Kam, K., Kalimuddin, S., Tan, S. Y., Loh, J., Thoon, K. C., Vasoo, S., Khong, W. X.,

Suhaimi, N.-A., Chan, S. J., Zhang, E., Oh, O., Ty, A., Tow, C., Chua, Y. X., Chaw, W. L., Ng, Y., Abdul-Rahman, F., Sahib, S., Zhao, Z., Tang, C., Low, C., Goh, E. H., Lim, G., Hou, Y., Roshan, I., Tan, J., Foo, K., Nandar, K., Kurupatham, L., Chan, P. P., Raj, P., Lin, Y., Said, Z., Lee, A., See, C., Markose, J., Tan, J., Chan, G., See, W., Peh, X., Cai, V., Chen, W. K., Li, Z., Soo, R., Chow, A. L., Wei, W., Farwin, A., and Ang, L. W. (2020). Investigation of three clusters of COVID-19 in singapore: implications for surveillance and response measures. *The Lancet*, 395(10229):1039–1046.

Qian, G. Q., Yang, N. B., Ding, F., Ma, A. H. Y., Wang, Z. Y., Shen, Y. F., Shi, C. W., Lian, X., Chu, J. G., Chen, L., Wang, Z. Y., Ren, D. W., Li, G. X., Chen, X. Q., Shen, H. J., and Chen, X. M. (2020). Epidemiologic and clinical characteristics of 91 hospitalized patients with COVID-19 in Zhejiang, China: A retrospective, multi-centre case series. *QJM: An International Journal of Medicine*, 113(7):474–481.

Qin, J., You, C., Lin, Q., Hu, T., Yu, S., and Zhou, X.-H. (2020). Estimation of incubation period distribution of COVID-19 using disease onset forward time: A novel cross-sectional and forward follow-up study. *Science Advances*, 6(33):eabc1202.

Quesada, J., López-Pineda, A., Gil-Guillén, V., Arriero-Marín, J., Gutiérrez, F., and Carratala-Munuera, C. (2021). Incubation period of COVID-19: A systematic review and meta-analysis. *Revista Clínica Española (English Edition)*, 221(2):109–117.

Rai, B., Shukla, A., and Dwivedi, L. K. (2021). Incubation period for COVID-19: A systematic review and meta-analysis. *Journal of Public Health*. Avaliable at:https://doi.org/10.1007/s10389-021-01478-1.

Ren, X., Li, Y., Yang, X., Li, Z., Cui, J., Zhu, A., Zhao, H., Yu, J., Nie, T., Ren, M., Dong, S., Cheng, Y., Chen, Q., Chang, Z., Sun, J., Wang, L., Feng, L., Gao, G. F., Feng, Z., and Li, Z. (2021). Evidence for pre-symptomatic transmission of coronavirus disease 2019 (COVID-19) in China. *Influenza and Other Respiratory Viruses*, 15(1):19–26.

Rodríguez-Cola, M., Jiménez-Velasco, I., Gutiérrez-Henares, F., López-Dolado, E., Gambarrutta-Malfatti, C., Vargas-Baquero, E., and Gil-Agudo, Á. (2020). Clinical features of coronavirus disease 2019 (COVID-19) in a cohort of patients with disability due to spinal cord injury. *Spinal Cord Series and Cases*, 6(1):1–6.

Ryu, S., Ali, S. T., Jang, C., Kim, B., and Cowling, B. J. (2020). Effect of nonpharmaceutical interventions on transmission of severe acute respiratory syndrome coronavirus 2, south korea, 2020. *Emerging Infectious Diseases*, 26(10):2406.

Sanche, S., Lin, Y. T., Xu, C., Romero-Severson, E., Hengartner, N., and Ke, R. (2020). High contagiousness and rapid spread of severe acute respiratory syndrome coronavirus 2. *Emerging Infectious Diseases*, 26(7):1470–1477.

Shen, Q., Guo, W., Guo, T., Li, J., He, W., Ni, S., Ouyang, X., Liu, J., Xie, Y., Tan, X., Zhou, Z., and Peng, H. (2020). Novel coronavirus infection in children outside of Wuhan, China. *Pediatric Pulmonology*, 55(6):1424–1429.

Shi, P., Gao, Y., Shen, Y., Chen, E., Chen, H., Liu, J., Chen, Y., Xiao, Y., Wang, K., Shi, C., and Lu, B. (2021). Characteristics and evaluation of the effectiveness of monitoring and control measures for the first 69 patients with COVID-19 from 18 January 2020 to 2 March in Wuxi, China. *Sustainable Cities and Society*, 64:102559.

Song, Q., Zhao, H., Fang, L., Liu, W., Zheng, C., and Zhang, Y. (2020). Study on assessing early epidemiological parameters of COVID-19 epidemic in China. *Chinese Epidemiology Journal*, 41(4):461–465.

Tan, W. Y. T., Wong, L. Y., Leo, Y. S., and Toh, M. P. H. S. (2020). Does incubation period of COVID-19 vary with age? A study of epidemiologically linked cases in singapore. *Epidemiology and Infection*, 148:e197.

Tian, H., Liu, Y., Li, Y., Wu, C.-H., Chen, B., Kraemer, M. U. G., Li, B., Cai, J., Xu, B., Yang, Q., Wang, B., Yang, P., Cui, Y., Song, Y., Zheng, P., Wang, Q., Bjornstad,

O. N., Yang, R., Grenfell, B. T., Pybus, O. G., and Dye, C. (2020a). An investigation of transmission control measures during the first 50 days of the COVID-19 epidemic in China. *Science*, 368(6491):638–642.

Tian, S., Hu, N., Lou, J., Chen, K., Kang, X., Xiang, Z., Chen, H., Wang, D., Liu, N., Liu, D., et al. (2020b). Characteristics of COVID-19 infection in Beijing. *Journal of Infection*, 80(4):401–406.

Tindale, L. C., Stockdale, J. E., Coombe, M., Garlock, E. S., Lau, W. Y. V., Saraswat, M., Zhang, L., Chen, D., Wallinga, J., and Colijn, C. (2020). Evidence for transmission of COVID-19 prior to symptom onset. *Elife*, 9:e57149.

Viego, V., Geri, M., Castiglia, J., and Jouglard, E. (2020). Incubation period and serial interval of COVID-19 in a chain of infections in Bahia Blanca (Argentina). *Ciência & Saúde Coletiva*, 25(9):3503–3510.

Wang, P., Lu, J., Jin, Y., Zhu, M., Wang, L., and Chen, S. (2020a). Epidemiological characteristics of 1212 COVID-19 patients in Henan, China. *medRxiv*.

Wang, X., Zhou, Q., He, Y., Liu, L., Ma, X., Wei, X., Jiang, N., Liang, L., Zheng, Y., Ma, L., Xu, Y., Yang, D., Zhang, J., Yang, B., Jiang, N., Deng, T., Zhai, B., Gao, Y., Liu, W., Bai, X., Pan, T., Wang, G., Chang, Y., Zhang, Z., Shi, H., Ma, W.-L., and Gao, Z. (2020b). Nosocomial outbreak of COVID-19 pneumonia in Wuhan, China. *European Respiratory Journal*, 55(6):2000544.

Wang, Y., Cao, Z., Zeng, D. D., Zhang, Q., and Luo, T. (2021). The collective wisdom in the COVID-19 research: Comparison and synthesis of epidemiological parameter estimates in preprints and peer-reviewed articles. *International Journal of Infectious Diseases*, 104:1–6.

Wassie, G. T., Azene, A. G., Bantie, G. M., Dessie, G., and Aragaw, A. M. (2020). Incubation period of severe acute respiratory syndrome novel coronavirus 2 that causes coronavirus disease 2019: A systematic review and meta-analysis. *Current Therapeutic Research*, 93:100607.

Wei, Y., Wei, L., Liu, Y., Huang, L., Shen, S., Zhang, R., Chen, J., Zhao, Y., Shen, H., and Chen, F. (2020). A systematic review and meta-analysis reveals long and dispersive incubation period of COVID-19. *medRxiv*.

Wen, Y., Wei, L., Li, Y., Tang, X., Feng, S., Leung, K., Wu, X., Pan, X.-F., Chen, C., Xia, J., Zou, X., Feng, T., and Mei, S. (2020). Epidemiological and clinical characteristics of coronavirus disease 2019 in Shenzhen, the largest migrant city of China. *medRxiv*.

Won, Y. S., Kim, J.-H., Ahn, C. Y., and Lee, H. (2021). Subcritical transmission in the early stage of COVID-19 in Korea. *International Journal of Environmental Research and Public Health*, 18(3):1265.

Wong, J., Chaw, L., Koh, W. C., Alikhan, M. F., Jamaludin, S. A., Poh, W. W. P., and Naing, L. (2020). Epidemiological investigation of the first 135 COVID-19 cases in brunei: Implications for surveillance, control, and travel restrictions. *The American Journal of Tropical Medicine and Hygiene*, 103(4):1608–1613.

Wu, J., Huang, Y., Tu, C., Bi, C., Chen, Z., Luo, L., Huang, M., Chen, M., Tan, C., Wang, Z., Wang, K., Liang, Y., Huang, J., Zheng, X., and Liu, J. (2020). Household transmission of SARS-CoV-2, Zhuhai, China, 2020. *Clinical Infectious Diseases*, 71(16):2099–2108.

Xia, W., Liao, J., Li, C., Li, Y., Qian, X., Sun, X., Xu, H., Mahai, G., Zhao, X., Shi, L., Liu, J., Yu, L., Wang, M., Wang, Q., Namat, A., Li, Y., Qu, J., Liu, Q., Lin, X., Cao, S., Huan, S., Xiao, J., Ruan, F., Wang, H., Xu, Q., Ding, X., Fang, X., Qiu, F., Ma, J., Zhang, Y., Wang, A., Xing, Y., and Xu, S. (2020). Transmission of corona virus disease 2019 during the incubation period may lead to a quarantine loophole. *medRxiv*.

Xiao, Z., Guo, W., Luo, Z., Liao, J., Wen, F., and Lin, Y. (2021). Examining geographical disparities in the incubation period of the COVID-19 infected cases in Shenzhen and Hefei, China. *Environmental Health and Preventive Medicine*, 26(1):1–10.

Xiao, Z., Xie, X., Guo, W., Luo, Z., Liao, J., Wen, F., Zhou, Q., Han, L., and Zheng, T. (2020). Examining the incubation period distributions of COVID-19 on Chinese patients with different travel histories. *The Journal of Infection in Developing Countries*, 14(04):323–327.

Xu, X.-W., Wu, X.-X., Jiang, X.-G., Xu, K.-J., Ying, L.-J., Ma, C.-L., Li, S.-B., Wang, H.-Y., Zhang, S., Gao, H.-N., Sheng, J.-F., Cai, H.-L., Qiu, Y.-Q., and Li, L.-J. (2020). Clinical findings in a group of patients infected with the 2019 novel coronavirus (SARS-CoV-2) outside of Wuhan, China: Retrospective case series. *BMJ*, 368:m606.

Yang, L., Dai, J., Zhao, J., Wang, Y., Deng, P., and Wang, J. (2020a). Estimation of incubation period and serial interval of COVID-19: analysis of 178 cases and 131 transmission chains in Hubei province, China. *Epidemiology and Infection*, 148:e117.

Yang, N., Shen, Y., Shi, C., Ma, A. H. Y., Zhang, X., Jian, X., Wang, L., Shi, J., Wu, C., Li, G., Fu, Y., Wang, K., Lu, M., and Qian, G. (2020b). In-flight transmission cluster of COVID-19: a retrospective case series. *Infectious Diseases*, 52(12):891–901.

Yang, X., Yu, Y., Xu, J., Shu, H., Xia, J., Liu, H., Wu, Y., Zhang, L., Yu, Z., Fang, M., Yu, T., Wang, Y., Pan, S., Zou, X., Yuan, S., and Shang, Y. (2020c). Clinical course and outcomes of critically ill patients with SARS-CoV-2 pneumonia in Wuhan, China: A single-centered, retrospective, observational study. *The Lancet Respiratory Medicine*, 8(5):475–481.

You, C., Deng, Y., Hu, W., Sun, J., Lin, Q., Zhou, F., Pang, C. H., Zhang, Y., Chen, Z., and Zhou, X.-H. (2020). Estimation of the time-varying reproduction number of COVID-19 outbreak in China. *International Journal of Hygiene and Environmental Health*, 228:113555.

Yu, X., Sun, X., Cui, P., Pan, H., Lin, S., Han, R., Jiang, C., Fang, Q., Kong, D., Zhu, Y., Zheng, Y., Gong, X., Xiao, W., Mao, S., Jin, B., Wu, H., and Fu, C. (2020). Epidemiological and clinical characteristics of 333 confirmed cases with coronavirus disease 2019 in Shanghai, China. *Transboundary and Emerging Diseases*, 67(4):1697–1707.

Zhang, J., Litvinova, M., Wang, W., Wang, Y., Deng, X., Chen, X., Li, M., Zheng, W., Yi, L., Chen, X., Wu, Q., Liang, Y., Wang, X., Yang, J., Sun, K., Longini, I. M., Halloran, M. E., Wu, P., Cowling, B. J., Merler, S., Viboud, C., Vespignani, A., Ajelli, M., and Yu, H. (2020a). Evolving epidemiology and transmission dynamics of coronavirus disease 2019 outside Hubei province, China: a descriptive and modelling study. *The Lancet Infectious Diseases*, 20(7):793–802.

Zhang, P., Wang, T., and Xie, S. X. (2020b). Meta-analysis of several epidemic characteristics of COVID-19. *Journal of Data Science*, 18(3):536–549.

Zhang, T., Ding, S., Zeng, Z., Cheng, H., Zhang, C., Mao, X., Pan, H., Xia, G., and Che, D. (2021). Estimation of incubation period and serial interval for SARS-CoV-2 in Jiangxi, China, and an updated meta-analysis. *The Journal of Infection in Developing Countries*, 15(03):326–332.

Zhao, C., Xu, Y., Zhang, X., Zhong, Y., Long, L., Zhan, W., Xu, T., Zhan, C., Chen, Y., Zhu, J., Xiao, W., and He, M. (2020). Public health initiatives from hospitalized patients with COVID-19, China. *Journal of Infection and Public Health*, 13(9):1229–1236.

Zhou, J., Xu, X.-P., Xu, F., Shao, Y., Zou, M.-H., Yu, J.-J., Liu, F., Zuo, W., Xie, S.-G., Zhou, C.-Y., and Zhang, W. (2020). Clinical symptoms and psychological changes of patients with COVID-19 in Jiangxi province. Available at: https://europepmc.org/article/PPR/PPR122471.
